# Cochlear Place Specificity of the Auditory Brainstem Response to Narrowband Chirp versus 2-1-2 stimuli: High Pass Noise/Derived Responses

**DOI:** 10.1101/2025.11.16.25340312

**Authors:** Ronald Nkansah Adjekum, David R. Stapells

## Abstract

**Objective:** In recent years, many researchers have recommended using narrowband chirp (NBchirp) stimuli for Auditory Brainstem Response (ABR) audiometry instead of more-standard 2-1-2 cycles linear-gated tones, primarily because NBchirps often result in larger ABR wave V amplitudes. However, the acoustic frequency spectra of currently recommended NBchirps are wider than those for 2-1-2 tones, and it is currently not known whether ABRs to these NBchirps have similar (or poorer) cochlear place specificity compared to 2-1-2 tones. The current study used the high-pass noise/derived response technique to assess the cochlear regions contributing to ABRs evoked by NBchirp versus 2-1-2 stimuli.

**Design:** A total of 24 adults with normal hearing participated (N=12 for each stimulus frequency). Stimuli were 60-dB peSPL 500- and 2000-Hz NBchirps and 2-1-2 tones mixed with high-pass (HP) filtered masking noise. The level of broadband (pink) noise required to mask the ABR was determined individually, then the broadband noise at this level was HP filtered at ½-octave intervals. Three ABR replications were obtained for each condition, with recordings stopped when the residual noise level of each replication was reduced to 40 nanovolts. Derived responses (DRs) representing 1-octave-wide or ½-octave-wide cochlear regions were calculated by subtracting ABRs recorded in HP noise.

**Results:** Non-masked ABR amplitudes in response to NBchirps were significantly larger than those to 2-1-2 stimuli, averaging 55% larger for 500 Hz and 81% larger for 2000 Hz. For both 500- and 2000-Hz stimuli, HP noise masking produced significant amplitude decreases, occurring 1 to ½ octave higher for ABRs to NBchirps versus 2-1-2 tones. One-octave-wide and ½-octave-wide DR amplitude profiles for the ABRs to 2-1-2 tones showed good cochlear place specificity, as described in previous studies. DR results for the NBchirps were similar but showed important differences. The profiles for the 2000-Hz NBchirps showed significantly larger amplitudes in the 4- and 1-kHz DRs compared to the 2-1-2 stimuli. Many more responses were seen 1-octave away for the 2000-Hz NBchirp compared to 2-1-2 tone. DR results for 500-Hz tones showed similar patterns but differences did not quite reach statistical significance, except amplitudes to NBchirps were larger at DR354, DR500 and DR707. A measure of the width of the 1-octave-wide and ½-octave-wide DR amplitude profiles (BW_0.075_, in Hz) showed the 500- and 2000-Hz NBchirp profiles were significantly wider (32% to 77%) compared to those for 2-1-2 stimuli. As the cochlear area able to respond decreased, wave V amplitudes to NBchirp stimuli decreased more than those for 2-1-2 stimuli, with no difference between stimuli for ½-octave-wide responses.

**Conclusion:** ABRs to narrowband chirps reflect wider cochlear contributions than those to 2-1-2 tones. Responses to NBchirps arise from cochlear regions as far as one octave away from the stimulus frequency. In contrast, responses to 2-1-2 tones arise from cochlear regions primarily within approximately ±0.5 octaves of the stimulus frequency. Further research in individuals with hearing loss is required to determine whether the wider bandwidths for NBchirps result in threshold mis-estimations, and whether NBchirp amplitude advantages over more-standard stimuli remain with hearing loss.

## INTRODUCTION

For several decades, researchers studying auditory brainstem response (ABR) audiometry have investigated the best choice of stimuli to estimate frequency-specific behavioral thresholds rapidly and accurately in patients who cannot be tested with conventional pure-tone audiometry. Many ABR studies have demonstrated that “2-1-2” tones (2 cycles rise, 1 cycle plateau, 2 cycles fall) and other brief tones with similar duration are good choices for predicting the behavioral pure-tone audiogram using the ABR (for review: Small & Stapells 2017). However, many have noted concerns about 2-1-2 and other similar brief tones, suggesting that ABR thresholds to these stimuli are too high and/or they lack frequency specificity (e.g., Scherg & Volk 1983; Sohmer & Kinarti 1984; Rodrigues et al. 2013; Laukli 2014; Ferm & Lightfoot 2015; Leusin Mattiazzi et al. 2024). In recent years, some have recommended using a newer stimulus, narrowband chirps (NBchirp), for threshold ABRs instead of 2-1-2 cycles linear-gated tones (Elberling & Don 2010), primarily because NBchirps often, though not always, result in larger ABR wave V amplitudes (Cobb & Stuart 2016; Ferm & Lightfoot 2015; Dzulkarnain et al. 2018; Pinto et al. 2022). NBchirps are brief tonal stimuli uprising in frequency which attempt to compensate for temporal dispersion along the cochlear partition by presenting the low-frequency components within the stimulus before the high-frequency components, according to cochlear travelling-wave delay models. Theoretically, if delay models are correct, aligning the arrival time of each frequency component in the stimulus to its place of maximum excitation along the basilar membrane should produce larger amplitude responses (Elberling et al. 2007; Gøtsche-Rasmussen et al. 2012).

NBchirps at four frequencies (500, 1000, 2000, and 4000 Hz) derived from the broadband CE-Chirp^®^ (level independent) developed by Elberling and colleagues are currently the most researched (and clinically utilized) NBchirp stimuli ^1^; they were constructed based on a delay model originally determined from derived-band ABR latencies (Elberling et al. 2007; Elberling & Don 2008). Based on the research cited above, these NBchirp stimuli are expected to elicit significantly larger ABR wave V amplitudes (compared to standard 5-cycle tonal stimuli). Nevertheless, it has also been argued that a “1-octave” wide region of the basilar membrane may be too small an area for the NBchirp to result in a significant increase in the ABR amplitude (Wegner & Dau 2002).

Most of the studies advocating the use of NBchirps have focussed on amplitude advantages over 2-1-2 stimuli, which would lead to improved signal-to-noise ratio, making ABR recordings faster and responses easier to detect (e.g., Bell et al. 2002; Ferm et al. 2013; Ferm & Lightfoot 2015; Cobb & Stuart 2016; Dzulkarnain et al. 2018; Talaat et al. 2019; Pinto et al. 2022). Nonetheless, the amplitude advantage of NBchirps is not consistently present for all stimulus frequencies and/or intensities (e.g., Bal 2022; Cobb & Stuart 2016; Dzulkarnain et al. 2018; Rodrigues et al. 2013; Megha et al. 2019; Ceylan et al. 2025). Some of the inconsistencies may be related to delay model issues. Most of these studies have been carried-out in adults with normal hearing; few have assessed NBchirp/standard tone ABR amplitude differences in either infants or individuals with hearing loss.

Previous studies have noted that the acoustic spectra of NBchirps appear wider than those of the 2-1-2 standard brief-tone stimuli (Bell et al. 2002; Cobb & Stuart 2016; Adjekum et al. 2024). Although current NBchirps are “octave band” stimuli (i.e., their spectrum is 1-octave-wide when measured at -3 dB), Adjekum and colleagues recently argued the bandwidth at -20 dB is a more realistic measure of the effective bandwidth of 2-1-2 or NBchirp spectra. Measured at -20 dB, NBchirp spectra bandwidths are 1.5 octaves, compared to 0.9 octaves for 2-1-2 stimuli (Adjekum et al. 2024). Because NBchirps have wider frequency spectra, it is likely the frequency and place specificity of ABRs to these stimuli will differ from 2-1-2 tones and may not be acceptable for clinical use. In other words, the ABRs to NBchirps may include contributions from unwanted frequency places (Bell et al. 2002; Adjekum et al. 2024). The wider spectral content of NBchirps compared to 2-1-2 stimuli may also explain the larger ABR amplitudes to these stimuli. However, stimulus spectrum is not always an accurate predictor of cochlear contributions to the ABR (e.g., linear- vs Blackman-windowed tones: Oates & Stapells 1997a; Oates & Stapells 1997b; Oates & Purdy 2001), thus it is currently not known whether ABRs to these NBchirps have worse (or similar) cochlear place specificity compared to 2-1-2 tones.

The high-pass noise/derived response (HPDR) technique has proven to be a useful method for quantifying cochlear contributions to an ABR to different stimuli. Teas, Eldridge, and Davis (1962) were the first to employ the HPDR subtraction technique to record auditory evoked potentials in animals. Later, Elberling (1974) and Eggermont (1976) employed this technique for electrocochleographic recordings in humans; Don and Eggermont (1978), as well as Parker and Thornton (1978) used this technique to assess the ABR in humans. The HPDR technique is applied by first recording ABRs in quiet and in the presence of broadband masking noise at an effective masking level, and then steeply high-pass (HP) filtering the noise at this intensity. The subtraction of responses recorded in HP noise with a lower cutoff frequency from responses recorded in HP noise with a higher cutoff frequency yield “derived responses” (DRs) that reflect the responses to stimulation of the basilar membrane approximately between the two cutoff frequencies (Don & Eggermont 1978; Parker & Thornton 1978; Picton et al. 1981; Evans & Elberling 1982; Stapells & Fok 2023). In a recent “masking within the derived band” study, Stapells and Fok (2023) confirmed the validity of the ABR HPDR method, with their results indicating 1-octave-wide separations in the HP noise settings resulted in responses reflecting 1-octave-wide bands with center frequencies closest to the lower HP cutoff in the HPDR calculation. The HPDR technique has proven to be an extremely valuable tool for investigation of mechanisms underlying the ABR, and, to a lesser extent, for clinical investigations. Most relevant to the current study, by assessing amplitudes of DRs representing narrow frequency bands in response to a stimulus, the HPDR technique provides a direct assessment of the cochlear contributions (i.e., “cochlear place specificity”) to the auditory evoked potential by that stimulus (Kramer 1992; Nousak & Stapells 1992; Oates & Stapells 1997b; Herdman et al. 2002; Stoll & Maddox 2024)

Several studies using the HPDR technique have shown good place specificity for the ABR to 2-1-2 tones and other 5-cycle tones (such as exact-Blackman windowed) (Kramer 1992; Nousak & Stapells 1992; Oates & Stapells 1997b). These studies have demonstrated ABR amplitude profiles showing (i) derived response (DR) response maxima at or within ±0.5 octave of the nominal stimulus frequency, indicating a place-specific response from the cochlea, (ii) sharp drop-offs above and below the DR frequency with maximum response amplitude, indicating little contribution from frequencies one or more octaves away from stimulus frequency, and (iii) center frequency and bandwidth measures which indicate place-specific contributions from the frequencies of interest. A similar assessment of cochlear place specificity has not been investigated for the ABR to NBchirps.

The current study used the HPDR method to investigate whether the acoustic spectral differences seen between NBchirp and 2-1-2 stimuli result in ABRs to the two stimuli showing differing cochlear contributions; specifically, whether the wider NBchirp spectra result in wider cochlear contributions to the ABR.

## MATERIALS AND METHODS

### Subjects

Twenty-four adults (N=12 per 500- and 2000-Hz stimulus frequency) with normal hearing were recruited for the study. The participants had no significant otologic or neurologic histories and were between the ages of 18 to 40 years. Subjects were assigned to groups randomly, without consideration of age or sex. Normal hearing required a pure-tone behavioral threshold of 15 dB HL (ANSI, 2023) or better at the octave frequencies of 250, 500, 1000, 2000, 4000, and 8000 Hz in the test ear. Additionally, normal middle-ear function was required, as demonstrated by a tympanogram with normal middle-ear pressure and normal tympanic membrane mobility (i.e., Type A tympanogram). For the ABR study, only one ear was tested (randomly selected between left and right ears).

### Stimuli and Calibration

The 2-1-2 and NBchirp stimuli were initially recorded from the research version of an Interacoustics Eclipse ABR system (EPx5 4.4 software). The Eclipse ABR system was set to deliver NB CE-Chirp^®^ LS and 2-1-2 stimuli at 100 dB pe SPL. The stimulus electrical output was fed directly to Sigview software (version 3.2) using a digitization rate of 44,100 Hz and a 95-ms analysis window. An average of 100 time-domain samples was obtained for each stimulus and then edited to include only the stimulus portion. These Sigview-recorded stimuli were then converted by an Intelligent Hearing Systems (IHS) SmartEP system to the SmartEP STM stimulus format (16-bit resolution at a rate of 40000 Hz). Figure 1 shows the acoustic spectra of these NBchirp and 2-1-2 stimuli. As previously demonstrated (Cobb & Stuart 2016; Adjekum et al. 2024), the NBchirp spectra are 70-80% wider (at -20 dB) than the 2-1-2 spectra. Comparisons of spectra of the stimuli presented by the IHS SmartEP and the Interacoustics Eclipse showed no difference between the stimuli (for both NB CE-Chirp^®^ LS and 2-1-2 stimuli).

**Figure 1.**
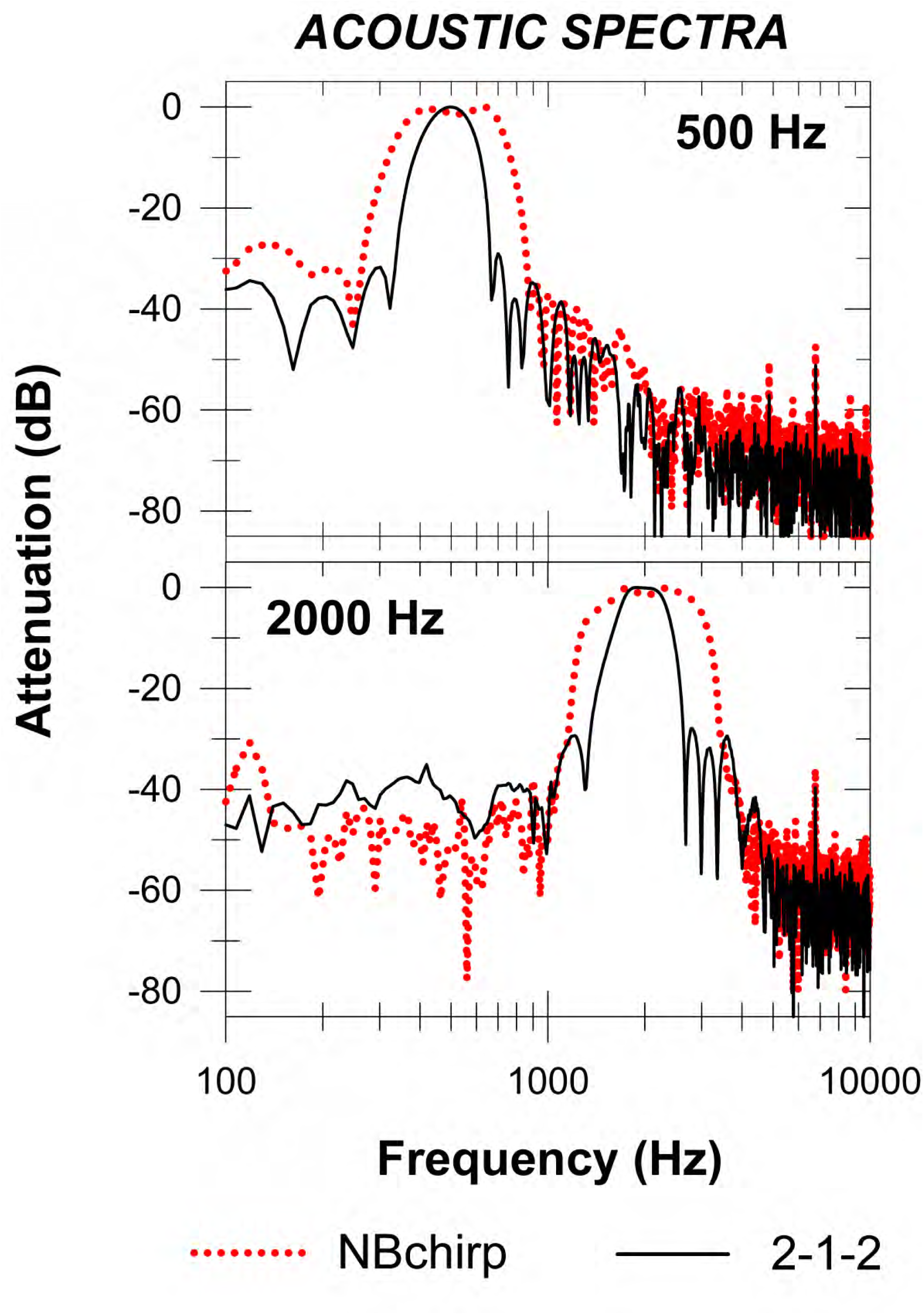
Acoustic spectra (output from ER-3A earphones) for NBchirp and 2-1-2 stimuli for 500- and 2000-Hz stimulus frequencies.

During ABR recordings, 500- and 2000-Hz stimuli (NBchirp and 2-1-2) were presented monaurally at 60 dB pe SPL via Etymotic Research ER-3A insert earphones. This level is equivalent to 40 (500 Hz) and 37 (2000 Hz) dB nHL for NBchirps (Gøtsche-Rasmussen et al. 2012)^2^ and to 38-41 (500 Hz) and 38-40 (2000 Hz) dB nHL for 2-1-2 stimuli (Fedtke & Richter 2007; Bagatto, 2020; Hatton et al. 2022). Stimuli were presented at a rate of 38.9/s with alternating stimulus polarity. Acoustic calibrations of the 60 dB pe SPL NBchirp and 2-1-2 stimuli were checked before each test using a GRAS RA0113 2-cc coupler and a 1-inch microphone (Larson Davis, Model 2575) and a PRM902 preamplifier with a Larsen Davis System 824 sound level meter.

### Broadband and High-Pass Noise Masking

Broadband pink noise (General Radio Company 1382 random-noise generator) was HP filtered using a cascade of two Stanford Research Systems (model SR650, each 115dB/octave slope) and then attenuated. HP noise cutoffs were selected to provide ½-octave separations. For 500-Hz stimuli, cutoffs were 8000, 4000, 2830, 2000, 1410, 1000, 707, 500, 354, and 250 Hz. For 2000-Hz stimuli, cutoffs were 16000, 8000, 5660, 4000, 2830, 2000, 1410, 1000, 707, and 500 Hz.^3^

Effective masker levels using broadband pink noise (HP filters both set at 0.001 Hz) were determined for each participant (and each stimulus). Initially, the levels of masking noise required to completely mask the stimuli behaviorally were obtained. Subsequently, levels required to mask the ABR were obtained starting at 5 dB above the behavioral masking level and then increasing the masker intensity using 1-dB steps until the ABR was determined to be masked, then a “down-2-dB-up-1-dB” bracketing method was used to establish the effective masking level/threshold. Once determined for a participant and stimulus, the effective masking level obtained for broadband pink noise was maintained for all ABR conditions with HP noise. Mean (±SD) behavioral masking levels for the 500-Hz stimuli were 64±2.1 dB SPL for NBchirps and 62±1.6 dB SPL for 2-1-2 stimuli. Behavioral masking levels for the 2000-Hz stimuli were 63 ±3.2 dB SPL for NBchirps and 62±1.3 dB SPL for 2-1-2 stimuli. Mean (±SD) ABR effective masking levels for 500-Hz stimuli were 77±2.5 dB SPL for NBchirps and 75±2.8 dB SPL for 2-1-2 stimuli. ABR effective masking levels for 2000-Hz stimuli were 76±2.7 dB SPL for NBchirps and 73±3.7 dB SPL for 2-1-2 stimuli.

All ABR recordings were obtained using the IHS SmartEP evoked potential system. Gold-plated electrodes were placed on the vertex (Cz) and mastoid (M1 or M2, ipsilateral to stimulated ear), with a low forehead (Fpz) electrode acting as ground. Interelectrode impedances were 3000 Ohms or less. The EEG recordings were amplified (100,000x), filtered (50-1500 Hz, 12 dB/octave), and digitized over a time window of 24.7 ms using a sampling rate of 20,000 Hz. An online residual noise (RN) calculation was calculated over a 10-ms window that included the wave V-V’ response but excluded stimulus artifact: 10.5-20.5 ms for 500 Hz and 6.5-16.5 ms for 2000 Hz. Individual recordings were continued until the RN was equivalent to ≤40 nanovolts (denoted as 0.080 µV by the SmartEP; Hatton et al. 2022).

Three replications of ABR waveforms, each with a RN ≤40 nV, when averaged together give an overall ABR waveform with a RN of approximately 23 nV. Because the derived-response subtraction procedure increases the noise in the ABR waveform by the square root of 2, the RN of the DRs (average of 3 replications) will be approximately 33 nV, which allows us to assess wave V-V’ responses with peak-to-peak amplitudes as low as 0.07 µV - 0.10 µV (i.e., 2-3x the RN) (e.g., Picton et al. 1983; Picton & Hink 1974; Don & Elberling 1996).

### Procedure

For most participants (22 of 24), this study involved one recording session of approximately 3 hours; 2 participants required two sessions to complete the study. Prior to conducting the experiments, ethics approval was obtained from our university’s Clinical Research Ethics Board. During ABR testing, participants were seated in a recliner chair and encouraged to relax and sleep. All testing was conducted in a double-walled sound-treated booth. Each participant was tested with the NBchirp and 2-1-2 stimuli in quiet, in the presence of broadband pink noise, and 10 HP masking noise conditions. Each participant was tested for only one stimulus frequency using both stimuli. The order of NBchirp versus 2-1-2 stimuli and HP noise cutoff frequencies was randomized for each participant.

### Derived Responses

Derived responses were obtained by subtracting, in succession, ABRs to the stimulus in HP noise at one cutoff frequency from the responses to the same stimulus in HP noise with a cutoff frequency either 1-octave or ½-octave higher. Thus, DRs for 1-octave- and ½-octave - wide analyses were obtained at each test frequency. The subtractions for the 1-octave and ½-octave DRs each resulted in 8 separate DRs, respectively. Calculations of the center frequencies of derived noise bands, formed by subtracting HP noise spectra using 1-octave and ½-octave HP noise separations, indicate that the center frequencies of these noise bands are closest to the frequency of the lower HP noise used in the DR subtractions.^4^ This is especially the case for ½-octave-wide noise bands. Recently, Stapells and Fok (2023) presented masked-ABR results which support the use of the lower HP noise cutoff to designate the center frequency of 1-octave-wide DRs. We therefore designated the lower frequency of the two HP noise cutoff frequencies used in the subtraction procedure as the DR center frequency, similar to previous studies by Stapells and colleagues (Picton et al. 1981; Nousak & Stapells 1992; Oates & Stapells 1997b; Herdman et al. 2002) and others (e.g., Don et al. 1979; Eggermont & Don 1980; Kramer 1992).

### Response Identification and Measurement

Three judges experienced in tone-ABR recordings determined the presence or absence of ABR Wave V, with agreement required by at least two judges. Results for the study showed reasonable agreement between the judges (Fleiss’ Kappa: 0.74), with response presence/absence agreement between all three judges of 81% across all conditions. ABR wave V amplitudes were measured for each subject for all conditions with present responses. Wave V amplitude was measured from wave V to the following wave V’ negativity. Wave V was defined as the maximum vertex-positive peak occurring between 6 and 20 ms after stimulus onset; in the case of multiple peaks of equal amplitude occurring between 6 and 20 ms, the peak that occurs before the largest negativity shift was selected. Wave V’ was defined as the largest negative point occurring within 8 ms following wave V (e.g., Stapells & Picton 1981; Oates & Stapells 1997a). If no response was present (as determined by the judges), then the waveform’s overall RN (for the average of all replications) was entered as the amplitude.

From the DR amplitude profiles (i.e., amplitude vs DR frequency), we also determined the bandwidth (in Hz) for a wave V amplitude at 0.075 µV (BW_0.075_), a level that represents a near-threshold response for both stimuli. Bandwidths were calculated by determining the frequency for a wave V-V’ amplitude of 0.075 µV on either side of the DR amplitude profile (using interpolation between frequencies), with the difference (in Hz) between the low and high frequencies calculated as bandwidth, and the geometric mean of these two frequencies calculated as the profile’s centre frequency. Supplemental Digital Content 2 provides a schematic example of one subject’s BW_0.075_ calculation.

### Statistical Analyses

The percentage of subjects showing present responses for NBchirp and 2-1-2 stimuli was calculated for each test condition. The percent of present ABRs for NBchirp versus 2-1-2 stimuli were compared using chi-square tests for DR results at ±1 octave away from the stimulus frequency.^5^ ABR wave V amplitudes to NBchirp and 2-1-2 stimuli in each condition (no-mask, HP Noise, 1-octave DR, and ½-octave DR) were summarized using means and standard deviations (SD). Wave V latencies were not assessed in the present study.

Dependent samples t-tests were calculated to analyze wave V amplitudes to NBchirp versus 2-1-2 tones in the no-mask condition. Repeated-measures ANOVAs were carried out on the HP noise and DR results. Two-way repeated-measures ANOVAs were calculated separately for the 500- and 2000-Hz stimuli to examine the effect of HP noise cutoff frequency on the amplitudes of ABR wave V-V’ to NBchirp versus 2-1-2 stimuli. Similarly, 2-way repeated-measures ANOVAs were computed separately for the 500- and 2000-Hz stimuli to analyze the effects of stimulus type and DR frequency on wave V amplitudes to NBchirp and 2-1-2 stimuli. One-octave-wide and ½-octave-wide DR amplitude results were analyzed separately. DR bandwidths were assessed using 2-way mixed-model ANOVAs (with stimulus frequency as a between factor and stimulus type as a within factor) were also calculated for the 500-and 2000-Hz 1-octave-wide bandwidths (in octaves), and for the ½-octave-wide DR bandwidths (in octaves) to NBchirp versus 2-1-2 stimuli. Finally, the effect of limiting cochlear responding bandwidth (CBW) on wave V amplitudes for the no-masking condition and the 1-octave-wide and ½-octave-wide DR conditions corresponding to the stimulus frequencies (i.e., DR500 and DR2000) were assessed using mixed-model ANOVA (STIMFRQ as a between-subject effect; STIM type and CBW as within-subjects effects), carried out for those participants with results for all three CBW conditions.

Where appropriate, Huynh-Feldt epsilon corrections were applied to the ANOVA degrees of freedom to compensate for any violations of sphericity (Huynh & Feldt 1976; Glass & Hopkins 1996). Neuman-Keuls *post hoc* analyses were applied where appropriate to analyze significant main effects and interactions in the ANOVA results (Glass & Hopkins 1996). With the exception of chi-square tests, results of all statistical testing were considered significant if p < .05. As there was a total of four chi-square tests carried-out on the percent-present results each for 1-octave and ½-octave wide DRs, after Bonferroni correction, significant chi-square results required p<.0125.

## RESULTS

### No-Mask Condition

The top of Figure 2A presents the grand-mean waveforms for the no-mask condition. Clear waves V are seen in the waveforms in the no-mask conditions in response to both stimuli for both frequencies. ABR waveforms for all participants for the No-Mask condition (as well as for the 1-octave-wide and ½-octave-wide DRs representing the stimulus frequency) are presented in Supplemental Digital Content 3. Mean (SD) wave V amplitudes to non-masked 500-Hz stimuli were 0.19 (0.06) µV for NBchirps and 0.13 (0.04) µV for 2-1-2 stimuli. Mean amplitudes to non-masked 2000-Hz stimuli were 0.27 (0.07) µV for NBchirps and 0.15 (0.04) µV for 2-1-2 stimuli. The larger amplitudes in the no-mask condition for NBchirp compared to 2-1-2 stimuli were statistically significant [500 Hz: *t*(11) = 3.8, p < .001; 2000 Hz: *t*(11) = 9.53, p < .001]. Almost all participants showed larger amplitudes to NBchirps (500 Hz: 11/12; 2000 Hz: 12/12) with mean NBchirp amplitude advantages being 55% for 500 Hz and 81% for 2000 Hz.

**Figure 2.**
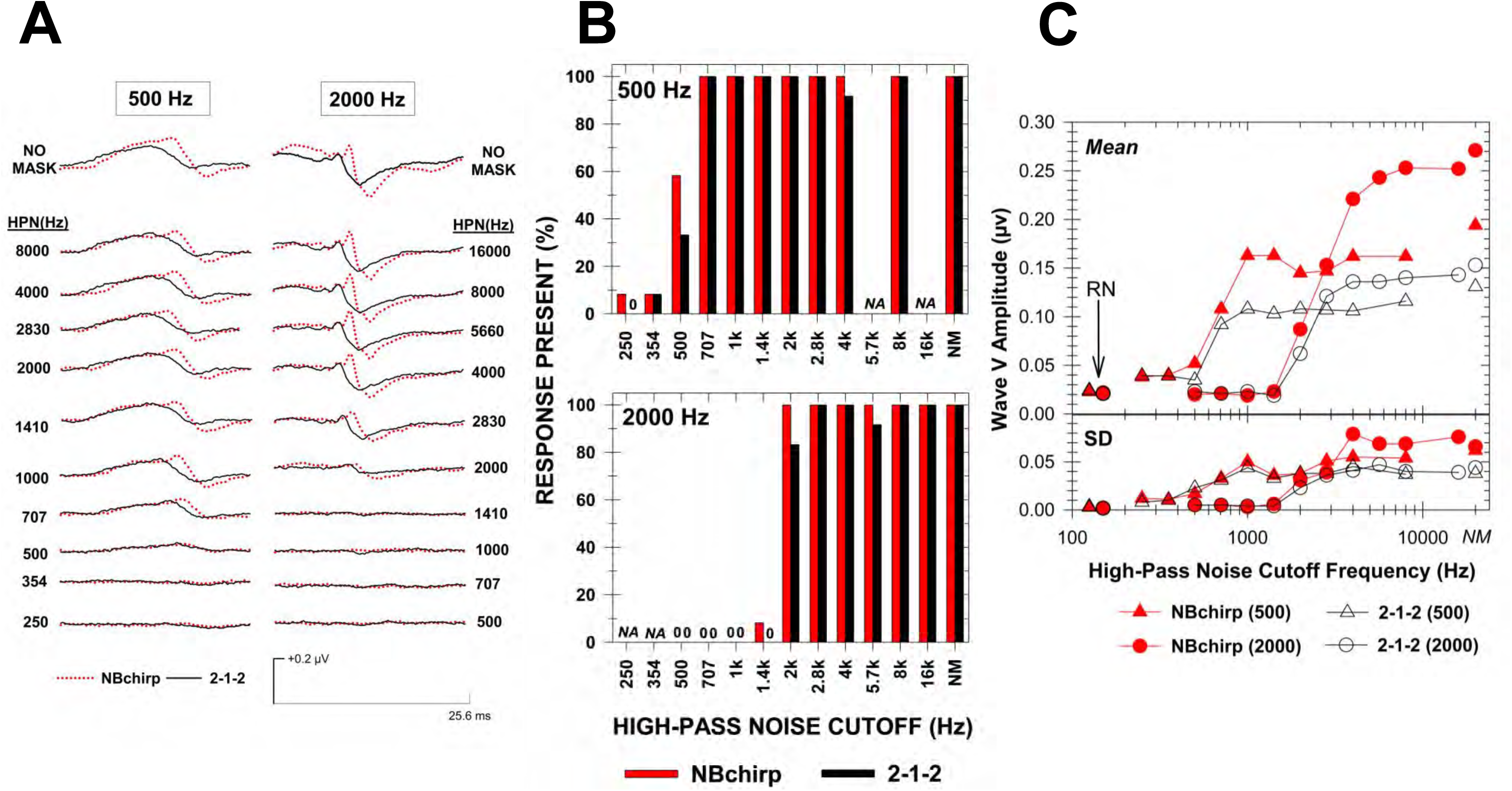
Results for No-Mask and HP noise conditions. N=12 participants per stimulus frequency. A, Grand-mean waveforms across all participants recorded to 60 dB pe SPL 500-Hz (left) and 2000-Hz (right) NBchirp and 2-1-2 stimuli for all 10 HP noise conditions, as well as the no-mask condition. B, Percent of participants showing “response present” ABR to NBchirp versus 2-1-2 stimuli in no-mask and HP noise conditions for 500-Hz (top) and 2000-Hz (bottom) stimuli. N=12 subjects per stimulus frequency. C, Mean and standard deviation (SD) amplitude results for the effects of HP noise cutoff on ABR wave V-V’ to NBchirp and 2-1-2 stimuli for 500- and 2000-Hz stimulus frequencies. The mean (and SD) residual noise (RN) across all HP conditions and participants is also plotted for each stimulus type and frequency. NM: no-mask condition; NA: not applicable (not tested).

### High-Pass Noise

Figure 2A also shows the grand-mean waveforms across all subjects to the 500- and 2000-Hz NBchirp and 2-1-2 stimuli for all 10 HP noise conditions. In general, the grand-mean waveforms indicated (i) larger responses to NBchirps, at least until the HP cutoff is within approximately ½ octave above the stimulus frequency, (ii) little change in responses until HP noise cutoff was within approximately 1 octave of the stimulus frequency, and (iii) large decrease in wave V amplitude once the HP noise cutoff was ½-octave above the stimulus frequency and below, with grand-mean waveforms showing little-or-no response once the HP noise cutoff is ½-octave below the stimulus frequency.

Figure 2B presents the percent of subjects showing present responses for each stimulus and HP noise condition. For responses to 500-Hz stimuli, response presence remained high (92-100%) for HP noise cutoffs down to 707 Hz, substantially decreasing (to 33-58%) for 500-Hz HP noise, and to 8% (N=1 subject with response) or less below. Similar results were seen for 2000-Hz stimuli, except that response presence did not substantially decrease until the HP noise cutoff was ½ octave *below* the stimulus frequency. No clear differences were seen in response presence between the two stimuli at either stimulus frequency.

Figure 2C presents mean (and SD) amplitude results for the effects of HP cutoff on responses to NBchirp and 2-1-2 stimuli for the two stimulus frequencies. In general, the amplitude profiles suggested (i) larger amplitudes to NBchirp vs 2-1-2 stimuli, (ii) larger responses to 2000-Hz vs 500-Hz stimuli (especially for NBchirps), and (iii) either no change or only small decreases in amplitude until the HP noise cutoff reaches approximately ½ to 1 octave above the stimulus frequency, with amplitudes then sharply decreasing below this. Two-way repeated-measures ANOVAs were carried-out separately for the 500-Hz and 2000-Hz stimulus frequencies. Results of these ANOVAs are shown at the top of Table 1.

**Table 1:** Results of two-way repeated measures ANOVAs comparing wave V amplitudes to NBchirp versus 2-1-2 stimuli in (i) HP noise conditions, (ii) 1-octave DR conditions and (iii) ½-octave DR conditions.

| Effect | 500-Hz stimuli |  |  |  |  | 2000-Hz stimuli |  |  |  |  |
| --- | --- | --- | --- | --- | --- | --- | --- | --- | --- | --- |
| | df | F | $\epsilon^\dagger$ | $p^\ddagger$ | $\eta_p^2$ | df | F | $\epsilon^\dagger$ | $p^\ddagger$ | $\eta_p^2$ |
| **HP NOISE** |  |  |  |  |  |  |  |  |  |  |
| HPFRQ | 10, 110 | 56.53 | 0.32 | <.001* | 0.84 | 10, 110 | 101.97 | 0.17 | <.001* | 0.90 |
| STIM | 1, 11 | 69.03 | 1.00 | <.001* | 0.86 | 1, 11 | 93.55 | 1.00 | <.001* | 0.90 |
| HPN x STIM | 10, 110 | 7.56 | 0.71 | <.001* | 0.41 | 10, 110 | 51.29 | 0.52 | <.001* | 0.82 |
| **1-OCT DR** |  |  |  |  |  |  |  |  |  |  |
| DRFRQ | 7, 77 | 56.34 | 0.65 | <.001* | 0.84 | 7, 77 | 58.43 | 0.41 | <.001* | 0.84 |
| STIM | 1, 11 | 1.46 | 1.00 | .253 | 0.12 | 1, 11 | 82.34 | 1.00 | <.001* | 0.88 |
| DRFRQ x STIM | 7, 77 | 3.86 | 0.68 | .005* | 0.26 | 7, 77 | 5.92 | 0.68 | <.001* | 0.35 |
| **½-OCT DR** |  |  |  |  |  |  |  |  |  |  |
| DRFRQ | 7, 70 | 32.78 | 0.82 | <.001* | 0.77 | 7, 70 | 26.79 | 0.77 | <.001* | 0.73 |
| STIM | 1, 10 | 3.70 | 1.00 | .083 | 0.27 | 1, 10 | 12.17 | 1.00 | .006* | 0.55 |
| DRFRQ x STIM | 7, 70 | 2.35 | 0.53 | .076 | 0.19 | 7, 70 | 4.91 | 1.00 | <.001* | 0.33 |

\* Significant ( $p < .05$ )
<sup>†</sup> Huynh–Feldt epsilon ( $\epsilon$ ) correction factor for degrees of freedom
<sup>‡</sup> Probability reflects corrected degrees of freedom
DR, Derived Response; HPN, High Pass Noise

The ANOVA results for each stimulus frequency indicated the significant main effect for HP noise cutoff frequency (HPFRQ) seen in the grand-mean waves and amplitude graphs. Similarly, the significant main effect for Stimulus Type (STIM) reflected the significantly larger amplitudes in response to NBchirps. However, the significant interaction (HPFRQ x STIM) indicated the above main effects may not hold across all conditions.

Neuman-Keuls *post hoc* analysis of the HPFRQ x STIM interaction for 500-Hz stimuli indicated: (i) amplitudes for NBchirp stimuli were significantly larger than 2-1-2 stimuli only for HP noise cutoffs of 1000 Hz and higher, (ii) amplitudes for NBchirp stimuli significantly decreased between 1000- and 707-Hz HP noise, and between 707- and 500-Hz HP noise cutoffs, whereas those for 2-1-2 stimuli only significantly decreased between 707- and 500-Hz cutoffs, and (iii) amplitudes for NBchirps significantly decreased from no-mask to HP noise 8000 Hz, a decrease which was not seen for 2-1-2 stimuli (with no significant decreases seen between 8000-through 1000-Hz HP noise cutoffs).

For the 2000-Hz stimuli, Neuman-Keuls *post hoc* analysis of the HPFRQ x STIM interaction indicated: (i) amplitudes for NBchirp stimuli were significantly larger than 2-1-2 stimuli only for HP noise cutoffs of 2000 Hz and higher (although statistically significant, the amplitude differences between stimuli were very small at HP2000 and HP2830), (ii) amplitudes for NBchirp stimuli significantly decreased between 5660- and 4000-Hz HP noise cutoffs, between 4000- and 2830-Hz HP noise cutoffs, between 2830- and 2000-Hz HP noise cutoffs, and between 2000- and 1410-Hz HP noise cutoffs, whereas those for the 2-1-2 stimuli only significantly decreased between 2830- and 2000-Hz HP noise cutoffs, and between 2000- and 1410-Hz HP noise cutoffs, and (iii) amplitudes for NBchirps significantly decreased from the no-mask to HP16000 conditions, a decrease which was not seen for 2-1-2 stimuli (with no significant decreases seen between 16000- through 5660-Hz HP noise cutoffs). Overall, the amplitude results with HP noise indicated responses to NBchirp stimuli were significantly reduced with HP noise cutoffs 0.5 to 1 octave higher than those seen for 2-1-2 stimuli.

### 1-Octave-Wide Derived Responses

Figure 3A shows grand-mean waveforms (across all subjects) in response to the 500- and 2000-Hz NBchirp and 2-1-2 stimuli for all eight 1-octave-wide derived-response conditions. [ABR waveforms for all participants for the 1-octave-wide DR500 and DR2000 conditions are presented in Supplemental Digital Content 3.] These grand-mean waveforms showed: (i) larger response amplitudes to NBchirps than to 2-1-2 stimuli, (ii) largest ABR amplitudes to both NBchirp and 2-1-2 stimuli in the DR bands within a ½ octave of the stimulus frequency, with amplitudes decreasing as DR frequency is further away from the stimulus frequency, (iii) for responses to the 2000-Hz tones, NBchirp results showed a clear wave V in the DR 1-octave above the stimulus frequency (i.e. DR4000), whereas there was no clear response for the 2-1-2 stimuli in the DR4000 waveform (both stimuli showed responses for DR1000), and (iv) for responses to the 500-Hz tones, differences between NBchirp and 2-1-2 stimuli were smaller and less-obvious, at least in the grand-mean waveforms. The grand-mean waveforms for 500-Hz stimuli showed small responses to both stimuli at 1-octave above (i.e., DR1000) but not 1-octave below (DR250) the stimulus frequency.

**Figure 3.**
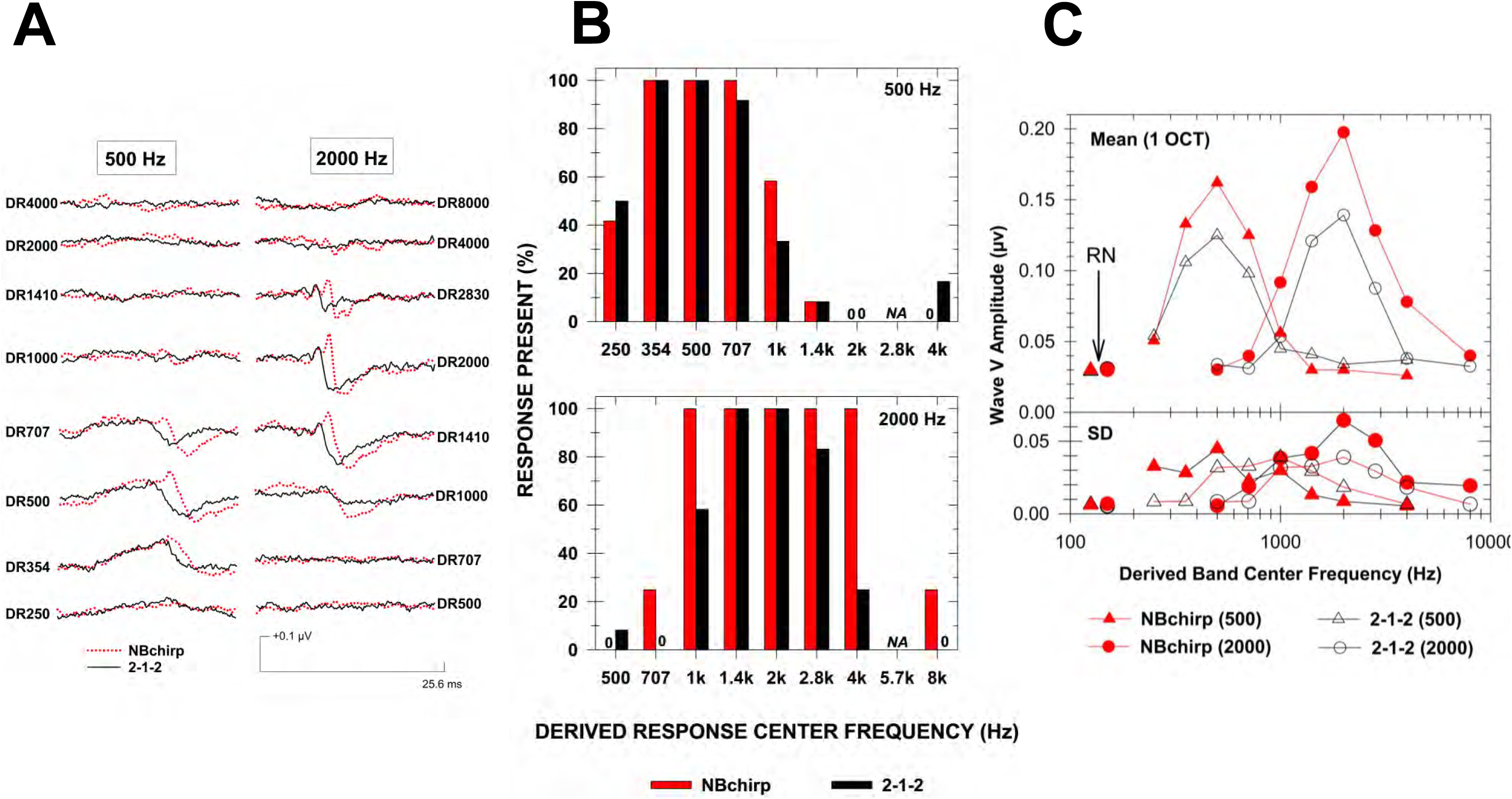
Results for 1-octave-wide DR conditions. N=12 participants per stimulus frequency. A, Grand-mean derived-response waveforms to 500-Hz (left) and 2000-Hz (right) NBchirp and 2-1-2 stimuli for all eight 1-octave-wide DR conditions. B, Percent of participants showing “response present” ABR to NBchirp versus 2-1-2 stimuli in 1-octave-wide DR conditions for 500-Hz (top) and 2000-Hz (bottom) stimuli. C, Mean and standard deviation (SD) amplitude results for the effects of 1-octave-wide DR center frequency on ABR wave V-V’ to NBchirp and 2-1-2 stimuli for 500- and 2000-Hz stimulus frequencies. The mean (and SD) residual noise (RN) across all DR conditions and participants is also plotted for each stimulus type and frequency. NA: not applicable (not tested).

Figure 3B presents the percent of subjects showing present responses to the NBchirp and 2-1-2 stimuli in each 1-octave-wide derived-response condition. For responses to the 500-Hz stimuli, response presence was 100% for the DR frequency representing stimulus frequency and remained high (92-100%) for DR conditions ½-octave away from the center frequency, then substantially decreased 1-octave away from the center frequency (DR250: 42% for NBchirp, 50% for 2-1-2; DR1000: 58% for NBchirp: 33% for 2-1-2). For responses to the 2000-Hz stimuli, similar to results for 500-Hz stimuli, response presence was 100% for the DR frequency representing stimulus frequency and remained high at DR conditions ½-octave away from the center frequency. In contrast to results for 500-Hz stimuli, however, at 1-octave away from the stimulus frequency (DR4000 and DR1000), DR results for 2000-Hz NBchirps continued to show 100% response presence, whereas DR results for 2000-Hz 2-1-2 stimuli showed substantially reduced response presence (DR1000: 100% for NBchirp, 58% for 2-1-2; DR4000: 100% for NBchirp: 25% for 2-1-2). The small differences in response presence one octave away for the 500-Hz stimuli did not reach statistical significance [DR250: χ^2^ (1, N=12) = 0.17, p=0.682; DR1000: χ^2^ (1, N=12) = 1.52, p=0.219]; however, differences for 2000-Hz stimuli were significant [DR1000: χ^2^ (1, N=12) = 6.32, p=0.0120; DR4000: χ^2^ (1, N=12) = 14.40, p<0.001], with NBchirps showing significantly more responses present.

Figure 3C presents mean (and SD) amplitude results for the effects of 1-octave-wide DR frequency on wave V amplitudes to NBchirp and 2-1-2 stimuli for the two stimulus frequencies. In general, the amplitude profiles showed: (i) both NBchirp and 2-1-2 amplitude profiles had largest mean amplitudes in the DR frequency representing the stimulus frequency, with mean amplitudes steeply decreasing in DRs above and below the stimulus frequency, especially when greater than ½-octave away, and (ii) NBchirp wave V amplitudes were greater than 2-1-2 amplitudes, especially within ½ octave of the stimulus frequency. Differences between stimuli decreased for DR frequencies ≥1-octave away from the stimulus frequency. Two-way repeated measures ANOVAs were carried-out separately for the 500- and 2000-Hz stimuli, with results shown in the middle of Table 1.

For 500-Hz stimuli, the significant main effect for DR frequency (DRFRQ) was consistent with the results seen in the grand-mean waves (Figure 3A) and amplitude profiles (Figure 3C): response amplitudes for both stimuli were significantly larger for the DR closest to the stimulus frequency and significantly decreased as DR frequency goes further away from 500 Hz. Although the main effect for stimulus type (STIM) was not significant, the significant DRFRQ x STIM interaction indicated significant differences existed between the two stimuli in some conditions. Neuman-Keuls *post hoc* analysis of this interaction for 500-Hz stimuli indicated significantly larger amplitudes in response to NBchirp compared to 2-1-2 stimuli for DR707, DR500, and DR354 conditions, with no differences between stimuli beyond these frequencies. Neuman-Keuls *post hoc* analysis also indicated response amplitudes for both stimuli significantly decreased for DR frequencies above 707 Hz and below 354 Hz.

For 2000-Hz stimuli, the significant main effect for DR frequency was also consistent with the results seen in the grand-mean waves (Figure 3A) and amplitude profiles (Figure 3C): response amplitudes were significantly larger for the DR closest to the stimulus frequency. As with the 500-Hz results, amplitude decreased significantly as the DR frequency went further away from 2000 Hz. The significant STIM (stimulus type) main effect indicated amplitudes of DRs to NBchirps were larger than those to 2-1-2 stimuli; however, the significant DRFRQxSTIM interaction indicated this was not be the case for all DR conditions. Neuman-Keuls *post-hoc* analysis of the significant DRFRQ x STIM interaction indicated the amplitudes in response to NBchirps were significantly larger than those to the 2-1-2 stimuli for DR4000, DR2830, DR2000, DR1410, and DR1000 Hz DR conditions. Similar to the results for 500-Hz stimuli, the Neuman-Keuls analyses for 2000-Hz stimuli also revealed that NBchirp amplitudes showed significant decreases starting ½-octave above and below the stimulus frequency, whereas amplitudes for 2-1-2 stimuli showed a significant decrease from DR2000 to DR1410 but only a non-significant trend (p < .1) to a smaller amplitude from DR2000 to DR2830.

The NBchirp amplitude profiles in Figure 3C appeared to be wider than those for the 2-1-2 amplitude profiles. Bandwidths (and center frequencies) of each subject’s 1-octave-wide amplitude profiles were determined for a wave V-V’ amplitude of 0.075 µV on either side of the profile (BW_0.075_). This was possible for all 12 subjects at 2000 Hz but not for one subject at 500 Hz, whose amplitudes for 2-1-2 stimuli did not allow us to determine a lower-side frequency for 0.075 µV. Bandwidth results for the 1-octave-wide DRs are shown at the top of Table 2. A mixed-model ANOVA (stimulus frequency as a between effect; stimulus type as a within effect) was calculated for the 500-and 2000-Hz 1-octave-wide DR bandwidths (converted to octaves) for NBchirp versus 2-1-2 stimuli. The ANOVA results indicated bandwidths of the NBchirp profiles were significantly wider than those for the 2-1-2 stimuli [STIM main effect: F(1, 21) = 51.75, p < .001, η^2^*_p_* = 0.71]. Expressed in octaves, there was no significant difference between the bandwidths of the profiles for 500- versus 2000-Hz stimuli [STIMFRQ main effect: F(1, 21) = 1.00, p = .328, η^2^*_p_* = 0.05]. There was a nonsignificant trend for NBchirp bandwidths, which were larger than 2-1-2 stimuli for both frequencies, to be even larger for 2000-Hz versus 500-Hz stimuli [STIMFRQ x STIM interaction: F(1, 21) = 3.71, p = .068, η^2^*_p_* = 0.15]. Also shown in Table 2, the center frequencies of the 500- and 2000-Hz 1-octave-wide DR amplitude profiles were very close to the stimulus frequency, with no obvious differences between NBchirp and 2-1-2 stimuli.

**Table 2:**
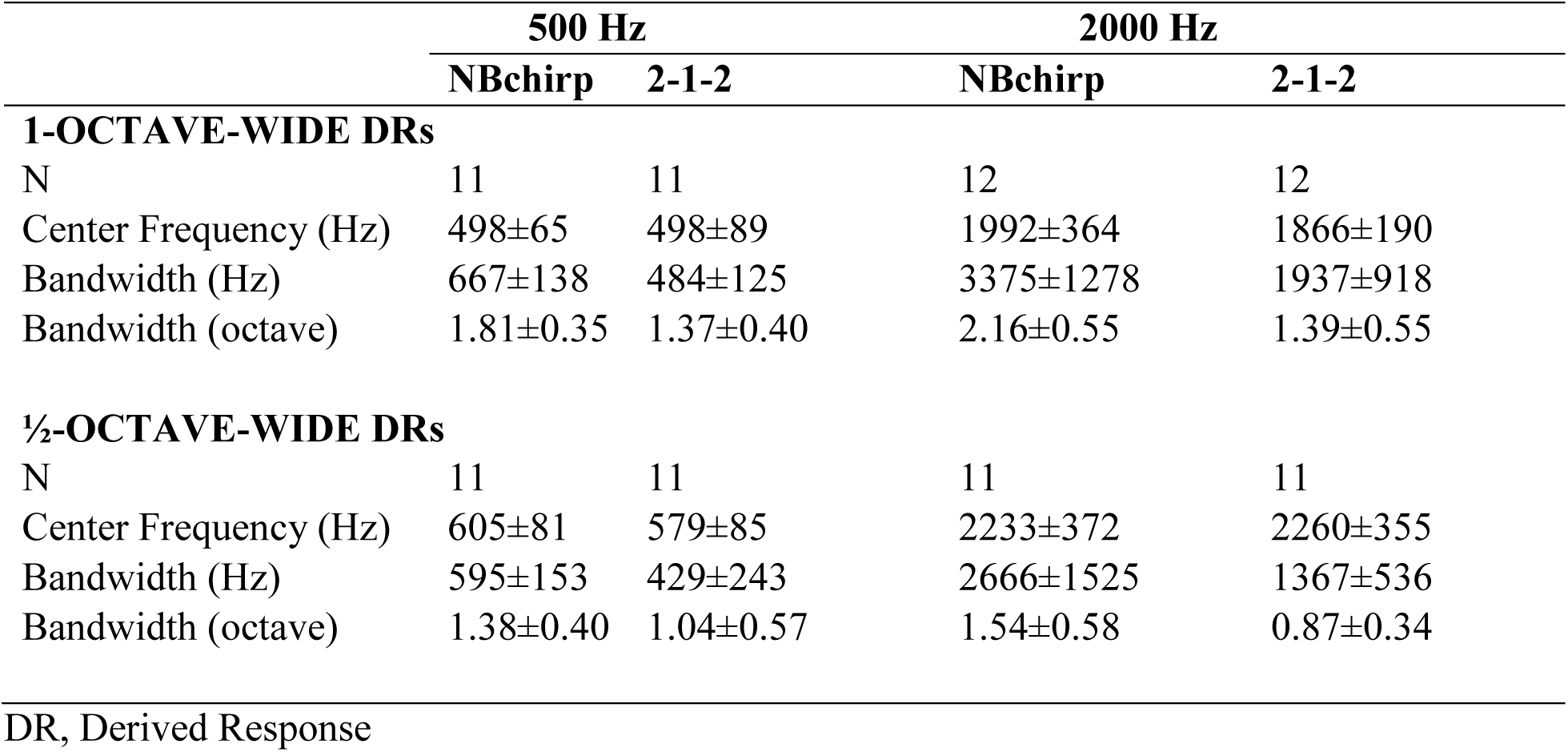
Mean (and standard deviation) center frequency and bandwidth (BW_0.075_) results for (i) 1-octave-wide and (ii) ½-octave-wide DR amplitude profiles to 500- and 2000-Hz NBchirp and 2-1-2 stimuli.

|  | 500 Hz |  | 2000 Hz |  |
| --- | --- | --- | --- | --- |
|  | NBchirp | 2-1-2 | NBchirp | 2-1-2 |
| **1-OCTAVE-WIDE DRs** |  |  |  |  |
| N | 11 | 11 | 12 | 12 |
| Center Frequency (Hz) | 498±65 | 498±89 | 1992±364 | 1866±190 |
| Bandwidth (Hz) | 667±138 | 484±125 | 3375±1278 | 1937±918 |
| Bandwidth (octave) | 1.81±0.35 | 1.37±0.40 | 2.16±0.55 | 1.39±0.55 |
| **$\frac{1}{2}$-OCTAVE-WIDE DRs** |  |  |  |  |
| N | 11 | 11 | 11 | 11 |
| Center Frequency (Hz) | 605±81 | 579±85 | 2233±372 | 2260±355 |
| Bandwidth (Hz) | 595±153 | 429±243 | 2666±1525 | 1367±536 |
| Bandwidth (octave) | 1.38±0.40 | 1.04±0.57 | 1.54±0.58 | 0.87±0.34 |

DR, Derived Response

### ½-Octave-Wide Derived Responses

To obtain a more frequency-specific evaluation of the contributions to the ABR by the 500- and 2000-Hz NBchirp and 2-1-2 stimuli, with better certainty of the center frequencies of derived bands (see Supplemental Digital Content 1), DRs representing ½-octave-wide bands were calculated. One subject in each of the 500- and 2000-Hz groups provided ½-octave DR results which were too difficult to interpret, likely due to the poorer signal-to-noise ratio of these responses. Thus, the sample size for the ½-octave results was 11 subjects per stimulus frequency. Figure 4A presents grand-mean waveforms to the 500- and 2000-Hz NBchirp and 2-1-2 stimuli for all eight ½-octave-wide DR conditions. [ABR waveforms for all participants for the ½-octave-wide DR500 and DR2000 conditions are presented in in Supplemental Digital Content 3.] The ½-octave waveforms appeared noisier because wave V amplitudes in the ½-octave-wide DRs were smaller compared to the 1-octave-wide DRs (note the scale difference between Figures 3A and 4A), which resulted in a smaller signal-to-noise ratio. The decrease in amplitude was due, at least in part, to the ½-octave-wide DRs originating from contributions of smaller portions of the cochlea. The residual electrical noise (RN) in the waveforms, however, was approximately the same between the ½- and 1-octave-wide DRs (on average, 31 vs 30 nV). In general, the grand-mean waveforms in Figure 4A were consistent with the observations of the grand-mean waveforms for the 1-octave-wide DR conditions (Figure 3A). Specifically, the grand-mean ½-octave DR waveforms indicated: (i) somewhat larger amplitudes in response to NBchirps than 2-1-2 stimuli, at least for 2000-Hz stimuli, (ii) the largest response amplitudes to both stimuli were within a half octave of the stimulus frequency, (iii) for the 2000-Hz stimuli, the grand-mean waveform to NBchirps showed a clear wave V response in the DR4000 grand mean (1-octave higher than the stimulus frequency) whereas there was no clear response to the 2-1-2 stimuli at this DR frequency, and (iv) for the 500-Hz stimuli, any differences in the pattern of response amplitudes between NBchirp versus 2-1-2 stimuli were relatively small, except the largest grand-mean NBchirp response was for DR707 versus DR500 for 2-1-2 stimuli.

**Figure 4.**
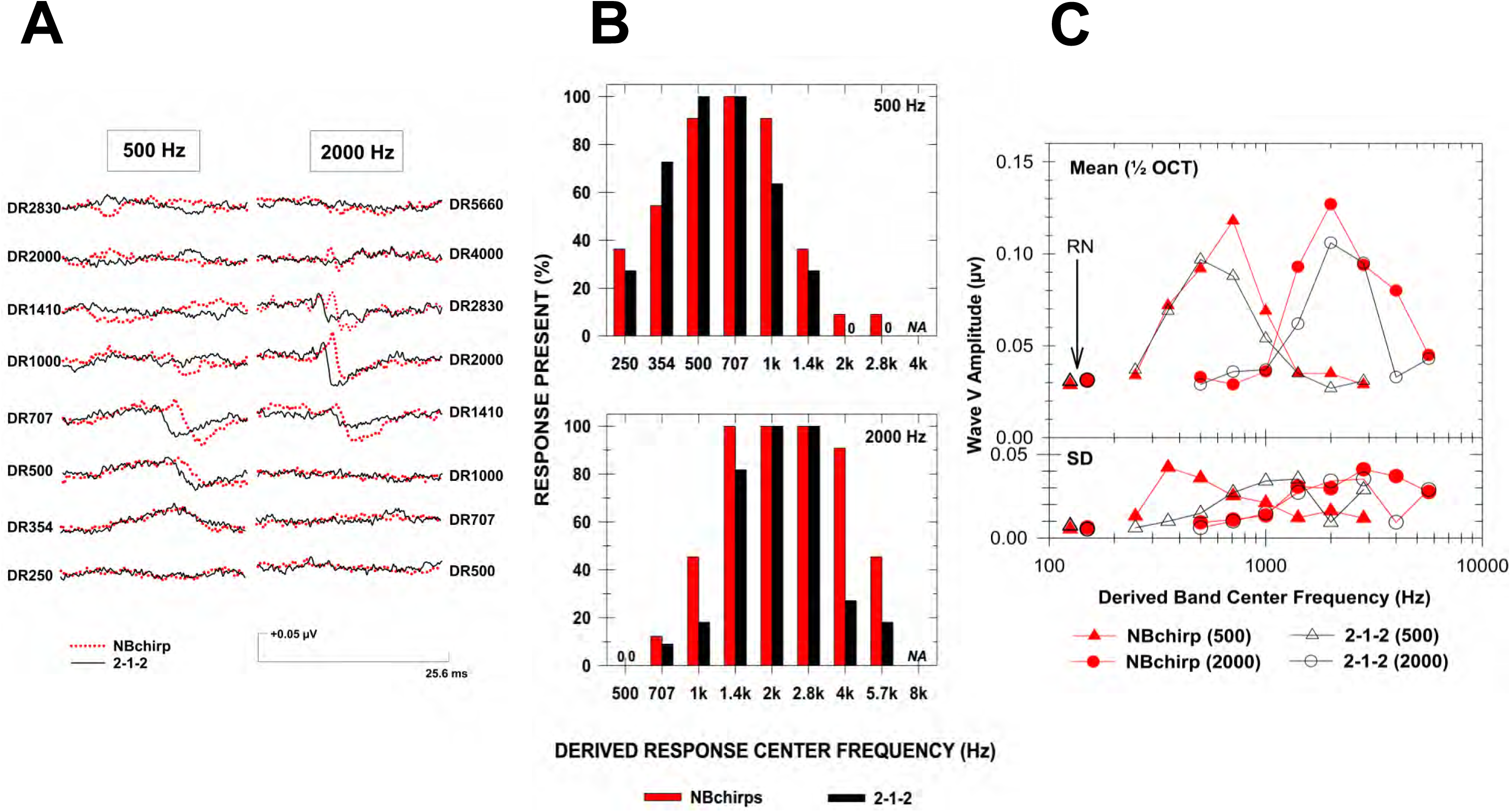
Results for ½-octave-wide DR conditions. N=11 participants per stimulus frequency. A, Grand-mean derived-response waveforms to 500-Hz (left) and 2000-Hz (right) NBchirp and 2-1-2 stimuli for all eight ½-octave -wide DR conditions. B, Percent of participants showing “response present” ABR to NBchirp versus 2-1-2 stimuli in the ½-octave-wide DR conditions for 500-Hz (top) and 2000-Hz (bottom) stimuli. C, Mean and standard deviation (SD) amplitude results for the effects of ½-octave-wide DR center frequency on ABR wave V-V’ to NBchirp and 2-1-2 stimuli for 500- and 2000-Hz stimulus frequencies. The mean (and SD) residual noise (RN) across all DR conditions and participants is also plotted for each stimulus type and frequency. NA: not applicable (not tested).

Figure 4B presents the percent of subjects showing present responses to the NBchirp and 2-1-2 stimuli at each ½-octave derived-response condition. For responses to 500-Hz stimuli, response presence was high (91-100%) at DR500 and DR707, with response presence decreasing 1-octave away and greater from the stimulus frequency. NBchirps response presence for DR1000 was higher than that for 2-1-2 stimuli (91% vs 64%); however, differences between response presence for NBchirp and 2-1-2 stimuli 1-octave away from 500 Hz did not reach statistical significance [DR250: χ^2^ (1, N=11) = 0.21, p=0.647; DR1000: χ^2^ (1, N=11) = 2.33, p=0.127]. For 2000-Hz stimuli, response presence for both stimuli was 100% at DR2000 and DR2830. At 1-octave above the stimulus frequency (DR4000), results for 2000-Hz NBchirps continued to show high (91%) response presence, whereas results for 2-1-2 stimuli showed substantially reduced response presence (27%). At 1-octave below the 2000-Hz stimulus frequency (DR1000), both stimuli showed low percent response presence (NBchirp: 46%; 2-1-2: 18%). The differences between NBchirp and 2-1-2 percent-present results did not reach statistical significance for DR1000 [χ^2^ (1, N=11) = 1.89, p=0.170], but 2000-Hz NBchirps showed significantly more responses for DR4000 [χ^2^ (1, N=11) = 9.21, p<0.001].

Figure 4C presents mean (and SD) amplitude results for the effects of DR frequency on ½-octave-wide DRs to NBchirp and 2-1-2 stimuli for the two frequencies. In general, the amplitude profiles showed: (i) generally, larger amplitudes to 2000-Hz tones for NBchirps compared to 2-1-2 stimuli, but only small differences between the stimuli in response to 500-Hz tones, (ii) largest mean amplitudes in the DRs within ½-octave of the stimulus frequency, with amplitudes substantially decreasing in DRs at least 1-octave away from the stimulus frequency, (iii) in response to 2000-Hz tones, relatively large mean amplitude to NBchirps in DR4000, whereas results for 2-1-2 stimuli in DR4000 showed very small mean amplitude (reflecting mostly “no-response” results for 2-1-2 stimuli at DR4000), and (iv) the amplitudes in response to NBchirps reflected the wider range of DR frequencies with responses than the 2-1-2 stimuli, consistent with the grand-mean waveforms (Figure 4A) and the response presence results (Figure 4B). Two-way repeated measures ANOVAs were conducted separately for amplitude results for the 500-Hz and 2000-Hz stimulus frequencies, with results shown in the bottom of Table 1.

For the ½-octave-wide DRs to 500-Hz stimuli, the significant main effect for DR frequency (DRFRQ) was consistent with the results seen in the grand-mean waves (Figure 4A) and amplitude profiles (Figure 4C). Both stimuli showed maximum responses within ½ octave of the stimulus frequency, with amplitudes significantly decreasing above and below. Neither the main effect for STIM nor the DRFRQxSTIM interaction reached statistical significance. The non-significant trends STIM and DRFRQxSTIM were likely due to the NBchirp mean amplitude being a little larger than that for 2-1-2 tones in the DR707 ½-octave -wide DR; otherwise, mean amplitudes were essentially the same in all other ½-octave -wide DRs, including DR500.

For the ½-octave -wide DRs to 2000-Hz stimuli, the two-way ANOVA showed the significant main effects of stimulus type (STIM), DR frequency (DRFRQ), as well as a significant DRFRQxSTIM interaction, which was consistent with the results seen in the grand-mean waves (Figure 4A), the percent-present histograms (Figure 4B), and the amplitude profiles (Figure 4C) for the ½-octave -wide DRs. Amplitudes decreased significantly as the DR frequency was further away from 2000 Hz. The STIM significant main effect of indicated significantly larger responses to NBchirps compared to 2-1-2 stimuli; however, this did not hold across all DR conditions. Neuman-Keuls *post hoc* analysis of the DRFRQxSTIM interaction indicated that the amplitudes in response to NBchirps were significantly larger than those to 2-1-2 stimuli in the DR4000, DR2000, and DR1410 conditions (but not in the DR2830 condition). Further, these analyses indicated that DR amplitudes to NBchirps significantly decreased between DR2000 and DR2830 but not between DR2830 and DR4000. In contrast, amplitudes in response to 2-1-2 stimuli did not significantly decrease between DR2000 and DR2830 but did significantly decrease between DR2830 and DR4000. Amplitudes to both stimuli significantly decreased when DR frequency was increased above DR4000 or below DR2000.

Similar to the 1-octave-wide DR results, the ½-octave -wide NBchirp amplitude profiles in Figure 4C appeared to be wider than those of the 2-1-2 amplitude profiles. Bandwidth (and center frequencies) of each subject’s ½-octave-wide DR amplitude profiles were determined for a wave V-V amplitude of 0.075 µV on either side of the profile. Summary results are shown in the bottom of Table 2. A mixed-model ANOVA (stimulus frequency as a between effect; stimulus type as a within effect) was carried-out for the 500-and 2000 Hz ½-octave-wide BW_0.075_ (in octaves) to NBchirp versus 2-1-2 stimuli. As with the 1-octave-wide results, the ANOVA indicated no significant differences in the width of the ½-octave-wide DR amplitude profiles between 500- and 2000-Hz stimuli when expressed in octaves [STIMFRQ main effect: F(1, 20) = 0.00, p = .987, η^2^*_p_* = 0.00]. However, the bandwidths of the ½-octave-wide DR NBchirp amplitude profiles were again significantly wider than those for the 2-1-2 stimuli [STIM main effect: F(1, 20) = 11.97, p = .003, η^2^*_p_* = 0.37], and this held for both stimulus frequencies, as indicated by the nonsignificant interaction [STIMFRQ x STIM interaction: F(1, 20) = 1.17, p = .293, η^2^*_p_* = 0.06]. As with the 1-octave-wide DR results, the ½-octave-wide DR amplitude profiles’ center frequencies were similar between the NBchirp and 2-1-2 stimuli.

### Effect of Cochlear Bandwidth: No-Mask, 1-Octave-Wide DR and ½-Octave-Wide DR

Insight into the effects of limiting the extent of the cochlear regions available to respond (cochlear bandwidth, CBW) to the NBchirp and 2-1-2 stimuli is provided by comparing wave V amplitudes in the no-mask condition with the 1-octave-wide and ½-octave-wide DR conditions with the same center frequency as the stimulus frequency (i.e., DR500 and DR2000), as shown in Table 3. A mixed-model ANOVA (STIMFRQ as a between effect; STIM type and CBW as within effects) was carried out for those participants with results for all three CBW conditions. The key finding was a significant CBW x STIM interaction [F(2, 40) = 38.87, *ε* = 1.00, p < .001, η^2^*_p_* = 0.66]. Wave V amplitudes in response to NBchirps compared to 2-1-2 stimuli, evaluated using Neuman-Keuls *post hoc* comparisons, showed greater decreases as cochlear bandwidth was narrowed from no-mask to 1-octave-wide to ½-octave wide DRs. Responses to 2-1-2 stimuli showed no significant change from no-mask to 1-octave-wide DRs, with a small albeit significant decrease in amplitude from 1-octave-wide to ½-octave-wide DRs. As already noted, wave V amplitudes in the no-mask condition were significantly larger in response to NBchirps compared to 2-1-2 stimuli; this amplitude advantage decreased in the 1-octave-wide condition, and was absent when responses are restricted to ½ octave around the stimulus frequency (i.e., there was no significant different between amplitudes to NBchirp versus 2-1-2 stimuli in the ½-octave-wide DR conditions corresponding to the stimulus frequencies). NBchirps showed larger responses to 2000-Hz stimuli, which was not seen with the 2-1-2 stimuli, as reflected by a significant STIM x STIMFRQ interaction [F(1, 20) = 5.01, *ε* = 1.00, p = .037, η^2^*_p_* = 0.20]. The CBW x STIM x STIMFRQ interaction was not significant [F(2, 40) = 1.13, *ε* = 1.00, p = .367, η^2^*_p_* = 0.05]. The difference in ½-octave-wide DR2000 amplitudes was not significant in this more-limited analysis compared to the significant Neuman-Keuls result reported above in the analysis which included all ½-octave-wide DR frequencies, but only results for 2000-Hz stimuli. Nevertheless, the difference between mean amplitudes was very small and of little practical significance (NBchirp: 0.13 µV; 2-1-2: 0.11µV).

**Table 3:** Effect of cochlear bandwidth on wave V amplitude (in µV and % of no-mask condition). Mean (±SD) results for 1-octave-wide and ½-octave-wide DR conditions with same CF as stimulus frequency (DR500 and DR2000). Results for those participants with complete data (N=11 per stimulus frequency).

| Stimulus<br>Frequency |  | Wide (No-Mask) |  | 1 Octave DR |  | Half Octave DR |  |
| --- | --- | --- | --- | --- | --- | --- | --- |
|  |  | NBchirp | 2-1-2 | NBchirp | 2-1-2 | NBchirp | 2-1-2 |
| 500 Hz | Amp in $\mu\text{V}$ | 0.20 | 0.14 | 0.16 | 0.13 | 0.09 | 0.10 |
| | ($\pm\text{SD}$) | $\pm 0.06$ | $\pm 0.04$ | $\pm 0.05$ | $\pm 0.05$ | $\pm 0.04$ | $\pm 0.02$ |
| | Amp<br>%No-Mask<br>($\pm\text{SD}$) | | | 82.4%<br>$\pm 16.0$ | 99.7%<br>$\pm 32.4$ | 48.0%<br>$\pm 25.0$ | 77.8%<br>$\pm 30.5$ |
| 2000 Hz | Amp in $\mu\text{V}$ | 0.28 | 0.16 | 0.21 | 0.15 | 0.13 | 0.11 |
| | ($\pm\text{SD}$) | $\pm 0.06$ | $\pm 0.04$ | $\pm 0.06$ | $\pm 0.03$ | $\pm 0.03$ | $\pm 0.03$ |
| | Amp<br>%No-Mask<br>($\pm\text{SD}$) | | | 74.0%<br>$\pm 11.9$ | 92.2%<br>$\pm 13.4$ | 45.8%<br>$\pm 8.7$ | 68.4%<br>$\pm 22.4$ |

DR, Derived Response; SD, Standard Deviation

The results also demonstrate another effect of restricting the cochlear bandwidth: although larger in amplitude measured in microvolts (at least, in the no-mask and some 1-octave DR conditions), relative to their non-masked amplitude, DR amplitudes (in percent-non-masked) for NBchirps decreased more compared to amplitudes for responses to 2-1-2 stimuli. A mixed-model ANOVA (STIMFRQ as a between effect; STIM type and CBW as within effects) was carried out on the percent-of-non-masked-amplitude results for those participants with results for both the 1-octave and ½-octave DR conditions. As shown in Table 3, expressed as a percent of non-masked amplitude, NBchirp DRs were significantly smaller than those for 2-1-2 stimuli [STIM main effect: F(1, 20) = 22.58, *ε* = 1.00, p < .001, η^2^*_p_* = 0.53]. Percent-amplitudes were also significantly smaller in the ½-octave DR condition compared to the 1-octave condition [CBW main effect: F(1, 20) = 55.52, *ε* = 1.00, p < .001, η^2^*_p_* = 0.74]. No other main effects or interactions were statistically significant [STIMFRQ: F(1, 20) = 1.07, *ε* = 1.00, p = .31, η^2^*_p_* = 0.05; CBW x STIMFRQ: F(1, 20) = 0.09, *ε* = 1.00, p = .76, η^2^*_p_* = 0.09; STIM x STIMFRQ: F(1, 20) = 0.11, *ε* = 1.00, p =.74, η^2^*_p_* = 0.01; CBW x STIM: F(1, 20) = 2.85, *ε* = 1.00, p =.11, η^2^*_p_* = 0.12; CBW x STIM x STIMFRQ: F(1, 20) = 0.65, *ε* = 1.00, p =.43, η^2^*_p_* = 0.03].

## DISCUSSION

Many studies have indicated larger ABR wave V (and Auditory Steady State Response, ASSR) amplitudes to NBchirp stimuli compared to more-standard stimuli. Though not consistently present for all frequencies and/or intensities, the larger amplitudes in response to NBchirp stimuli have been primarily attributed to the timing of frequencies within these stimuli based on cochlear travelling wave delays. However, the larger amplitudes seen with NBchirps might simply be due to these stimuli having wider spectra (Figure 1) and, thus, wider cochlear contributions. The current study is the first study to directly assess cochlear contributions to ABR wave V to current NBchirps using the high-pass noise/derived response (HPDR) technique. Results showed significantly wider cochlear contributions to ABR wave V to NBchirp compared to 2-1-2 stimuli.

### ABR to NBchirp and 2-1-2 Stimuli in High-Pass Noise

ABR wave V-V’ amplitudes in response to non-masked 60 dB pe SPL 500- and 2000-Hz NBchirps were significantly larger than those to 2-1-2 stimuli and almost all participants (23/24) showed larger responses to non-masked NBchirps. This finding is consistent with some previous studies showing significantly larger wave V amplitudes to 500- and 2000-Hz NBchirps compared to standard 5-cycle stimuli. However, the literature is quite inconsistent: some studies have shown larger wave V amplitudes to NBchirps at low (≤ 40 dB nHL) stimulus levels (e.g., Ferm et al. 2013; Rodrigues et al. 2013; Ferm & Lightfoot 2015). Other studies, however, have reported nonsignificant differences in the amplitude of responses to NBchirp and 2-1-2 stimuli at low stimulus levels (e.g., Dzulkarnain et al. 2018; Pinto et al. 2022; Ceylan et al. 2025).

Additionally, some reported no differences to higher stimulus levels (e.g., Megha et al. 2019; Bal 2022) and some have reported no differences (or responses to standard tones larger) specifically to 500-Hz stimuli (e.g., Rodrigues et al. 2013; Cobb & Stuart 2016). There is a large diversity across these studies in study subject populations (adults, older children, infants) and, especially, ABR recording parameters, which may explain some of the inconsistency. Despite this, it seems most studies have concluded wave V amplitudes are larger in response to NBchirps. This study’s HP noise and DR results provided some insight concerning these response amplitudes.

In the current study, averaged across all HP noise conditions, NBchirp stimuli produced larger responses than 2-1-2 stimuli. However, the amplitude difference between responses substantially decreased once the HP noise cutoff was within ½-octave above the stimulus frequency. Thus, the large gain in amplitude by NBchirps over 2-1-2 stimuli required contributions from cochlear frequency regions more than ½-octave higher than the stimulus frequency.

The effects of HP noise cutoff frequency on wave V amplitudes showed differences between NBchirp and 2-1-2 stimuli in the frequency at which HP noise cutoffs produced significant ABR amplitude changes. ABR amplitudes to NBchirp stimuli were significantly reduced with HP noise cutoffs one-half octave (500-Hz stimuli) to 1-octave (2000-Hz stimuli) higher than those noise cutoffs which resulted in significant decreases in ABR amplitude for 2-1-2stimuli. This suggests that the ABR to NBchirps receive contributions from wider cochlear frequency places than the responses to 2-1-2 stimuli, at least at 60 dB pe SPL.

In the current study, the HP filtered noise had a very steep slope but only 40-45 dB stopband attenuation. The effects of HP masking suggested some leakage of noise occurred into frequencies below the HP filter cutoffs. Noise leakage effects were primarily decreases in response amplitude in the 16000-Hz and/or 8000-Hz HP noise conditions. Similar findings with HP masking of the ABR have previously been reported and discussed (e.g., Oates & Stapells 1997a). As 16,000 Hz is well above the output limits of the ER-3A earphones, any response changes are likely due to noise leakage below the 16,000-Hz cutoff (noise at 73-77 dB SPL with a 50-dB stopband attenuation could still see 23-27 dB SPL noise leaking into the stopband). Given it is unlikely that 60 dB peSPL 2000-Hz stimuli (NBchirp or 2-1-2) would produce responses from such high-frequency cochlear regions (nor, indeed, from 8000-Hz regions for 500-Hz stimuli), the more likely explanation is noise leakage into the filter stopband. However, significant amplitude decreases in the 16000- or 8000-Hz HP noise conditions were only observed for NBchirps and not for 2-1-2 stimuli. One would expect leakage to affect responses to both stimuli equally. ABRs to NBchirps originating from wider frequency regions might explain this finding.

### Derived Responses to NBchirp versus 2-1-2 Stimuli

Cochlear place specificity of ABR wave V to NBchirps and 2-1-2 stimuli is demonstrated by the amplitude profiles (graph of amplitude vs DR frequency) of the 1-octave-wide and ½-octave-wide DRs. In general, these amplitude profiles showed that responses to both NBchirp and 2-1-2 stimuli originated primarily from cochlear contributions representing frequencies within one octave of the stimulus frequency. Thus, both stimuli demonstrate cochlear place specificity. More-detailed assessment of the results, however, suggested significantly better cochlear place specificity for the ABR to 2-1-2 stimuli compared to NBchirps.

The DR grand-mean waveforms, percent of subjects showing present responses at each DR condition, and DR amplitude profiles, center frequencies, and bandwidth measures all indicated good place specificity of the ABR to the 500- and 2000-Hz 2-1-2 stimuli. Specifically, these measures showed: (i) response presence limited primarily to DRs within ± 0.5 octave of the stimulus frequency, (ii) response amplitude maxima at the nominal stimulus frequency or within a half octave of the nominal stimulus frequency, (iii) a sharp drop-off in the amplitude of responses approximately one-half to 1-octave above and below the DR frequency with the maximum response amplitude (usually at the stimulus frequency), indicating that the cochlear contributions to the ABR wave V to the 2-1-2 stimuli are primarily within approximately within ±0.5 octaves of the stimulus frequency, and (iv) the mean center frequencies calculated for the 1-octave-and ½-octave-wide DR amplitude profiles to the 2-1-2 stimuli were close to the nominal stimulus frequencies of the 2-1-2 stimuli. NBchirp amplitude profiles were similar in shape to those for 2-1-2 stimuli, with center frequencies close to the stimulus frequency. However, NBchirps showed more responses present at frequencies one octave higher (and sometimes one octave lower) as well as significantly larger amplitudes one octave above and below the stimulus frequency.

A measure of the width (in Hz) of the amplitude profiles provided an estimate of the bandwidth of the responses to each stimulus and, thus, an easy comparison between stimuli. In the current study, we determined bandwidth from the amplitude profiles for a wave V amplitude of 0.075 µV, a level that represents a near-threshold response for both stimuli. Mean ½-octave-wide DR bandwidths obtained for the ABRs to 2-1-2 stimuli in the current study were 1.04 octaves for 500 Hz and 0.87 octaves for 2000 Hz, thus approximating a 1-octave-wide basilar membrane place. In comparison, at 1.38 octaves for 500 Hz and 1.54 octaves for 2000 Hz, mean ½-octave-wide DR bandwidths for the NBchirp amplitude profiles were significantly wider than those for 2-1-2 stimuli, and well over 1-octave-wide. Bandwidths measured from the 1-octave-wide DR amplitude profiles were also significantly wider for NBchirps compared to 2-1-2 stimuli. Thus, NBchirps produce larger contributions from a wider cochlear region compared to 2-1-2 stimuli.

In previous studies using a variety of masking methods, the bandwidths for ABRs to more-standard 500- and 2000-Hz tones have demonstrated largest cochlear contributions originate from frequency places within approximately ±0.5 octave of the stimulus nominal frequency (e.g., Klein 1983; Horiuchi 1989; Wu & Stapells 1994; Abdala & Folsom 1995; Oates & Stapells 1997b; Oates & Purdy 2001). Using the HPDR technique, Oates showed wave V DR (½-octave-wide) amplitude profile bandwidths of 622 and 2058 Hz for 500- and 2000-Hz 2-1-2 stimuli (at 80 dB peSPL), respectively (Oates 1996). In the current study, ½-octave-wide DR bandwidths for ABRs to 2-1-2 stimuli (428 and 1367 Hz) were much smaller than those reported by Oates, likely due to the lower stimulus intensity employed in the current study (60 pe SPL).

Although larger in amplitude measured in microvolts, DR amplitudes for NBchirps decreased more relative to their non-masked amplitude than DR amplitudes for 2-1-2 stimuli as cochlear bandwidth was decreased. The significant differences in the DR percent amplitudes between NBchirp and 2-1-2 stimuli likely cannot be attributed to differences in noise masking levels, as approximately the same levels (within 2-3 dB) of masking noise were used for masking the ABR to the NBchirp and 2-1-2 stimuli. A more likely explanation for the NBchirp DR percent amplitudes being significantly lower than those for 2-1-2 stimuli is that the responses to non-masked NBchirps originate from a wider cochlear frequency range (as demonstrated by this study’s HP noise and DR results), thus response contributions to the non-masked ABR are spread out over more DRs, and individual DRs are smaller expressed as a percentage of the no-mask response amplitude.

### NBchirp Amplitude Advantage versus Cochlear Bandwidth

The ABR wave V-V’ amplitudes in response to non-masked 60 dB pe SPL 500- and 2000-Hz NBchirps were significantly larger than those to 2-1-2 stimuli. This amplitude advantage for NBchirps has been shown by many studies, and is the primary reason for recommending NBchirps over more-standard tones, although it is not consistently present in all intensity/frequency conditions (see above). However, as the results of the current study clearly demonstrate, the NBchirp amplitude advantage requires a response from a wider cochlear range. When cochlear bandwidth was reduced by the HPDR technique to 1 octave, the advantage remained but was significantly decreased. This counters the suggestion by Wegner and Dau (2002) that a 1-octave wide bandwidth is not sufficient to produce a chirp amplitude advantage. When the cochlear bandwidth was limited to a ½-octave-wide region, the NBchirp amplitude advantage was essentially absent.

The results of the current study cannot unequivocally determine whether the larger amplitude of responses to non-masked NBchirps is due to their wider acoustic spectrum (Figure 1) or to the optimized timing of frequencies within the NBchirp to overlap responses from different frequency regions such that they constructively superimpose and result in a larger response. Reducing the cochlear bandwidth of the response using the HPDR technique would effectively limit and perhaps eliminate the effects of the optimized timing, especially for the ½-octave-wide DR condition. Even if the effects of optimized timing are absent, ½-octave-wide DR results indicate NBchirps produce responses from a wider cochlear region than 2-1-2 stimuli. Similar results were seen when HP noise cutoffs were within 0.5-1 octave above the stimulus frequency. Thus, as cochlear bandwidth decreased (from wideband to 1-octave-wide to ½-octave-wide), there was less cochlear “space” for NBchirps to utilize any amplitude advantage of dispersing frequencies according to travelling wave delays. From these results, one could postulate that the ½-octave-wide DRs for NBchirps reflected primarily spectral contributions rather than spectral *plus* optimization of frequency timing, and thus, little-or-no difference existed between response amplitudes to the two stimuli.

### Clinical Implications

As already noted, many previous studies have assessed the cochlear place specificity of the ABR to more-standard tonal stimuli, such as 2-1-2 linear-gated tones and 5-cycle Blackman-windowed tones. Previous research has employed pure-tone masking, HP noise masking, and notched-noise masking techniques (e.g., Klein 1983; Horiuchi 1989; Wu & Stapells 1994; Abdala & Folsom 1995; Oates & Stapells 1997a; Oates & Stapells 1997b; Oates & Purdy 2001). Others have studied 2-1-2 and similar stimuli in individuals with steeply sloping cochlear hearing loss (Davis et al. 1984; Purdy & Abbas 2002). In general, results from these studies suggest the ABR to standard tones such as 2-1-2 stimuli show reasonably good cochlear place specificity, at least up to about 80 dB pe SPL. However, when audiogram configurations show particularly steep hearing losses (≥50-60 dB/octave slope), standard tones may misestimate hearing thresholds for the frequencies with elevated thresholds (Davis et al. 1984; Horiuchi 1989; Stapells et al. 1990; Purdy & Abbas 2002).

To our knowledge, no previous studies have assessed the place specificity of the ABR to NBchirps by systematically varying masker frequencies. A number of studies, however, have assessed NBchirps in individuals with sensorineural hearing loss using the ABR and/or ASSR (e.g., Seidel et al. 2015; Lee et al. 2016; Sininger et al. 2018; Talaat et al. 2019; Eder et al. 2020; Ehrmann-Müller et al. 2021; Leusin Mattiazzi et al. 2024; Khan & Vanaja 2025; Galhoum et al. 2023; Michel & Jørgensen 2017), with most showing reasonably good threshold estimation results for NBchirps. NBchirp threshold results for several individual cases with mostly gently or moderately sloping hearing loss (15-35 dB/octave slope) plus 1 case with steeply sloping loss have also been published (Seidel et al. 2015; Mühler et al. 2012; Khan & Vanaja 2025). However, none of these studies have assessed NBchirp results in groups of individuals with steeply/precipitously sloping audiogram configurations (≥40 dB/octave slope). Testing ABR thresholds in only individuals with flat or mildly sloping audiograms does not adequately assess cochlear place specificity (Davis et al. 1984; Purdy & Abbas 2002). Indeed, only two studies have directly compared ABR to NBchirps vs standard tones in individuals with hearing loss (Talaat et al. 2019; Khan & Vanaja 2025), none in infants. Results of these two studies indicate good threshold estimates using either NBchirp or standard tone stimuli, with results from Talaat and colleagues also suggesting faster test times and apparently larger amplitudes for NBchirps, though results details were not provided (Talaat et al. 2019). Importantly, based on the mean audiograms presented by these 2 studies, most individuals with hearing loss had flat or gently sloping audiogram configurations and thus would not provide a test of any frequency or place specificity differences between stimuli (Talaat et al. 2019; Khan & Vanaja 2025). Due to NBchirp’s wider cochlear activation, as indicated by the results of the present study, ABR thresholds for NBchirp stimuli may be more likely to misestimate the severity of hearing thresholds, compared to 2-1-2 stimuli. Misestimation of thresholds would most likely be an issue primarily when there are large differences in thresholds between adjacent octave (and especially half-octave) audiometric frequencies; that is, steeply falling (or falling) audiograms. Further research is needed comparing NBchirp and 2-1-2 stimuli ABR thresholds in individuals, especially infants and young children, with steeply sloping hearing loss.

As suggested above, the ABR amplitude advantage for NBchirps over standard tones is the primary motivation for using NBchirps for ABR audiometry. This advantage, however, has not been shown to be consistently present for all stimulus frequency/intensity conditions. Moreover, as the current study shows, as the available response bandwidth narrows (to 1 octave or to ½ octave), the amplitude advantage decreases and even disappears. It is currently an open question whether sensorineural hearing losses remove/reduce this amplitude advantage (perhaps due to the audiogram configuration and/or to cochlear traveling wave properties which differ from those used in the model to produce the NBchirps). Clearly, more research is needed comparing ABR results (amplitudes and thresholds) to NBchirp and 2-1-2 stimuli in individuals with hearing loss.

How wide of a spectrum is acceptable for ABR stimuli is a longstanding matter of debate within Audiology (Davis 1976; Stapells & Picton 1981; Davis et al. 1984; Sohmer & Kinarti 1984; Laukli & Mair 1986; Gorga & Thornton 1989; Laukli 2014; McCreery et al. 2015). In cases with normal hearing or hearing loss with flat or mildly sloping/rising configurations, ABR thresholds to 2-1-2 or NBchirp stimuli, for the most part, represent the pure-tone behavioral thresholds at the stimulus frequencies. When hearing losses show a steeper configuration, this is not always the case (Davis et al. 1984; Horiuchi 1989; Stapells et al. 1990; Purdy & Abbas 2002). The ±0.5-octave cochlear place bandwidth of the ABR to 2-1-2 tones seems to be accepted within the field, given most early hearing programs use either standard brief tones (such as 2-1-2 linear-gated or 5-cycle Blackman-windowed tones) or NBchirps (which are usually described as being “octave-band”). Based on the current study, the ABR to 2-1-2 stimuli may reflect contributions from cochlear places representing frequencies within one-half octave of the stimulus frequency (e.g., from 1414 to 2828 Hz in response to 2000-Hz stimuli). On the other hand, the much wider bandwidth of ABRs to NBchirps – as wide as 2 octaves (Table 2) -- would seem to be too wide. As this study demonstrated, the 2000-Hz NBchirp produced responses from the 4000-Hz cochlear region in most subjects (compared to few for 2-1-2 stimuli). Concern about NBchirp frequency specificity has led some researchers to suggest embedding NBchirp stimuli in HP noise or notched noise (Corona-Strauss et al. 2012; Baljić et al. 2017b). However, as the current study shows, restricting the responding cochlear region using masking noise reduces (or eliminates) the wave V amplitude advantage of NBchirps over 2-1-2 stimuli. Should future research show reduced (or absent) amplitude advantages for NBchirps in individuals with hearing loss, there would seem little reason to using NBchirp stimuli with such a wide bandwidth. Further research in individuals with hearing loss is needed.

### Study Limitations

The current study investigated cochlear place specificity for a relatively low stimulus intensity (60 dB pe SPL, approximately 40 dB nHL). Although the frequency and place specificity of the ABR to standard tonal stimuli at higher stimulus intensities has previously been studied (e.g., Klein 1983; Horiuchi 1989; Oates & Stapells 1997b; Oates & Purdy 2001), studies are clearly needed on the place specificity of the ABR to higher intensity NBchirps (e.g., 80-90 dB pe SPL). Additionally, this study only investigated the place specificity of the ABR to 500-and 2000-Hz NBchirps. There is a need to conduct further studies on the place specificity of the ABR to 1000- and 4000-Hz NBchirps. For example, it may be that the place specificity of the ABR to 4000-Hz NBchirps will be different from that of the ABR to 2000-Hz NBchirps because of the reduction in the frequency content of the 4000-Hz NBchirp by the ER-3A earphone. The current study showed smaller differences between NBchirp vs 2-1-2 results for 500-Hz stimuli; it is not known whether results for 1000-Hz NBchirps would be similar to the place specificity results seen in the present study for 2000-Hz vs 500-Hz stimuli.

This study only assessed results from 12 normal-hearing participants for each stimulus frequency. This relatively small sample size may have reduced this study’s ability to detect small differences between stimuli and conditions (for example, response presence differences 1 octave away from stimulus frequency).

The current study assessed results only from adult participants with normal hearing. Although these results are likely to be applicable (at least partially) to individuals with cochlear hearing loss, research in individuals (especially infants) with hearing loss is required. The NBchirp delay models are derived from travelling wave data of the normal adult cochlea, and these delay models may not represent delays found in infant and/or pathological cochleae. This could result in suboptimal amplitude enhancement or even response cancellation (i.e., response amplitudes to NBchirps no different from, or even smaller than, those to 2-1-2 stimuli). Importantly, further studies are needed to investigate whether individuals with steep (rising or falling) cochlear hearing loss configurations show differences in amplitudes or thresholds for ABR thresholds to NBchirp and 2-1-2 stimuli.

### Conclusions

Although both stimuli demonstrate cochlear place specificity, ABRs to NBchirps show significantly poorer place specificity compared to ABRs to 2-1-2 stimuli. As the relative amplitude advantage for NBchirps over 2-1-2 stimuli decreases as the region of the cochlea contributing to the response decreases, it is not yet clear whether NBchirps amplitude advantages over more-standard stimuli exist when there is cochlear hearing loss. Further research, especially in individuals with steep cochlear hearing loss, is required.

## Supporting information

Supplemental Digital Content #1, #2, #3

## Data Availability

ABR amplitude data produced in the present study are available upon reasonable request to the authors

## Ethical approval

Ethics approval was obtained from the University of British Columbia Clinical Research Ethics Board (#H20-02458; P.I. Dr. N. Shahnaz)

## Financial disclosures/conflicts of interest

This study was supported by a Natural Sciences and Engineering Research Council of Canada Discovery Grant to Susan Small (RGPIN-2018-04655). Financial support for R.N.A. was provided by: (i) a Four-Year Doctoral Fellowship from the University of British Columbia (UBC), and (ii) the UBC School of Audiology and Speech Sciences. There are no conflicts of interest, financial, or otherwise.

## Author contribution statement

R.N.A. and D.R.S. designed the experimental procedures and analyses. R.N.A. recruited participants and ran the experiments, analyzed the ABR waveforms and managed results’ databases. R.N.A. and D.R.S. analyzed and discussed the research findings and wrote the article content.

## Acknowledgements

This study was carried-out in Dr. Susan Small’s laboratory. A strong supporter of this research, due to her illness and subsequent death, Dr. Small was unable to participate in the planning or completion of this study. This study was supported by a Natural Sciences and Engineering Research Council of Canada Discovery Grant to Susan Small (RGPIN-2018-04655). Financial support for R.N.A. was provided by: (i) a Four-Year Doctoral Fellowship from the University of British Columbia (UBC), and (ii) the UBC School of Audiology and Speech Sciences. An earlier version of this article has been posted on the medrXiv preprint server: https://www.medrxiv.org/content/10.1101/2025.11.16.25340312v1

## Footnotes

1 The current NB CE-Chirp^®^ LS narrowband chirps are based on the level-independent broadband CE-Chirp^®^ (as opposed to the level-specific broadband CE-Chirp^®^ LS). NB CE-Chirp^®^ LS narrowband chirps are level-specific only in their onset timings are adjusted such that wave V latencies should be about the same independent of intensity. NB CE-Chirp^®^ LS narrowband chirp durations and spectra are not affected by intensity (Adjekum et al. 2024).

2 After correction for the use of different couplers: IEC 60126 2-cc vs IEC 60711 (Haughton 2006),

3 The ER-3A earphone response output is limited to frequencies below about 10,000 Hz (Elberling et al. 2012; Etymotic Research, 2025). ER-2 earphones have wider bandwidth and were initially considered for this study. However, ER-3A earphones were finally chosen as they are the earphones most used clinically. The “HP16000” HP noise condition was kept for the study with the possibility it might provide additional insight concerning HP noise filter characteristics.

4 The calculations of center frequencies of the derived (i.e., subtracted) noise bands, as well as the nominal and measured HP noise cutoffs, are presented in Supplemental Data Content 1.

5 Observing responses within ±0.5 octave of the stimulus frequency would likely not be a concern for audiometry. However, obtaining responses as far as 1-octave away from the stimulus frequency would be very problematic clinically. Beyond 1-octave away, there were too few responses for either stimulus.

## Notes

### Competing Interest Statement

The authors have declared no competing interest.

### Author Declarations

The Clinical Research Ethics Board of the University of British Columbia gave ethical approval for this work.

### Summary of Updates

Figures have been revised (as well as any Figure callouts in text).

