## Supplemental Digital Content #1, #2, #3 for "Cochlear Place Specificity of the Auditory Brainstem Response to Narrowband Chirp versus 2-1-2 stimuli: High Pass Noise/Derived Responses"

#### List of Supplemental Digital Content:

Supplemental Digital Content 1 [SDC-1-Noise&DerivedBandMeasures(Nov8-2025).pdf]

Supplemental Digital Content 2 [SDC-2-SampleCalculation of Bandwidth(Oct28-2025).pdf]

Supplemental Digital Content 3 [SDC-3-Individual waves(Nov14-2025).pdf]

AUTHORS' ORIGINAL MANUSCRIPT

#### High-Pass Noise: Nominal and measured (@ -3 dB) High-Pass Noise Cutoffs (electric)

**Table SDC1.1: Nominal and measured high-pass noise cutoffs**

| **Nominal (Hz)** | | **250** | **354** | **500** | **707** | **1000** | **1410** | **2000** | **2830** | **4000** | **5660** | **8000** | **16000** |
|---|---|---|---|---|---|---|---|---|---|---|---|---|---|
| **Measured (Hz)** | | 238 | 330 | 469 | 667 | 947 | 1312 | 1878 | 2652 | 3762 | 5343 | 7563 | 15147 |

After cascading of the two SR650 HP filters; Measured at -3 dB from max;
Stopband slope: 43-47 dB per half-octave (equivalent to 86-94 dB per octave).
Measured stopband attenuation: 40-45 dB

#### Measured center frequencies (CFs) of 1-octave and ½-octave derived bands

Broadband pink noise the center frequency was measured as the geometric mean of the upper and lower -20 dB cutoff points and the bandwidth was estimated as the difference between the upper and lower -20 dB points.

#### ONE-OCTAVE-WIDE DERIVED BANDS

##### Table SDC1.2: One-octave-wide derived bands measured from high-pass noise subtractions (electric)

| **NOMINAL CENTER FREQUENCY (Hz)** | **250** | **354** | **500** | **707** | **1000** | **1410** | **2000** | **2830** | **4000** |
|---|---|---|---|---|---|---|---|---|---|
| HP nominal cutoffs (Hz) | 250-500 | 354-707 | 500-1000 | 707-1410 | 1000-2000 | 1410-2830 | 2000-4000 | 2830-5660 | 4000-8000 |
| **MEASURED CENTER FREQUENCY (Hz)** | **303** | **426** | **606** | **861** | **1225** | **1735** | **2429** | **3486** | **4934** |
| Nominal Geometric Mean (Hz) | 354 | 500 | 707 | 1000 | 1410 | 2000 | 2830 | 4000 | 5660 |
| Lower bound @-20dB (Hz) | 221 | 308 | 440 | 625 | 895 | 1257 | 1784 | 2527 | 3554 |
| Upper bound @-20dB (Hz) | 413 | 589 | 836 | 1187 | 1677 | 2394 | 3307 | 4810 | 6848 |
| Bandwidth @-20dB (Hz) | 192 | 281 | 397 | 563 | 78 | 1137 | 1523 | 2283 | 3294 |
| Bandwidth (octave) | 0.90 | 0.93 | 0.93 | 0.93 | 0.91 | 0.93 | 0.89 | 0.93 | 0.95 |

AUTHORS' ORIGINAL MANUSCRIPT

#### Table SDC1.3: One-octave-wide derived bands measured from high-pass noise subtractions (acoustic)

| **NOMINAL CENTER FREQUENCY (Hz)** | **250** | **354** | **500** | **707** | **1000** | **1410** | **2000** | **2830** | **4000** |
|---|---|---|---|---|---|---|---|---|---|
| HP nominal cutoffs (Hz) | 250-500 | 354-707 | 500-1000 | 707-1410 | 1000-2000 | 1410-2830 | 2000-4000 | 2830-5660 | 4000-8000 |
| **MEASURED CENTER FREQUENCY (Hz)** | **309** | **439** | **621** | **879** | **1251** | **1748** | **2479** | **3519** | **4386** |
| Nominal Geometric Mean (Hz) | 354 | 500 | 707 | 1000 | 1410 | 2000 | 2830 | 4000 | 5660 |
| Lower bound @-20dB (Hz) | 221 | 312 | 444 | 626 | 909 | 1251 | 1763 | 2535 | 3545 |
| Upper bound @-20dB (Hz) | 432 | 619 | 870 | 1233 | 1722 | 2442 | 3485 | 4886 | 5428 |
| Bandwidth @-20dB (Hz) | 212 | 307 | 426 | 607 | 812 | 1190 | 1722 | 2352 | 1883 |
| Bandwidth (octave) | 0.97 | 0.99 | 0.97 | 0.98 | 0.92 | 0.96 | 0.98 | 0.95 | 0.61 |

#### HALF-OCTAVE-WIDE DERIVED BANDS

#### Table SDC1.4: Half-octave-wide derived bands measured from high-pass noise subtractions (electric)

| **NOMINAL CENTER FREQUENCY (Hz)** | **250** | **354** | **500** | **707** | **1000** | **1410** | **2000** | **2830** | **4000** | **5660** |
|---|---|---|---|---|---|---|---|---|---|---|
| HP nominal cutoffs (Hz) | 250-354 | 354-500 | 500-707 | 707-1000 | 1000-1410 | 1410-2000 | 2000-2830 | 2830-4000 | 4000-5660 | 5660-8000 |
| **MEASURED CENTER FREQUENCY (Hz)** | **255** | **361** | **510** | **727** | **1030** | **1462** | **2067** | **2927** | **4141** | **5880** |
| Nominal Geometric Mean (Hz) | 297 | 421 | 595 | 841 | 1187 | 1679 | 2379 | 3365 | 4758 | 6729 |
| Lower bound @-20dB (Hz) | 220 | 31 | 436 | 623 | 896 | 1246 | 1800 | 2513 | 3514 | 4997 |
| Upper bound @-20dB (Hz) | 296 | 416 | 597 | 847 | 1184 | 1715 | 2370 | 3410 | 4881 | 6919 |
| Bandwidth @-20dB (Hz) | 76 | 102 | 161 | 224 | 289 | 469 | 570 | 897 | 1367 | 1922 |
| Bandwidth (octave) | 0.43 | 0.40 | 0.45 | 0.44 | 0.40 | 0.46 | 0.40 | 0.44 | 0.47 | 0.47 |

AUTHORS' ORIGINAL MANUSCRIPT

#### Table SDC1.5: Half-octave-wide derived bands measured from high-pass noise subtractions (acoustic)

| **NOMINAL CENTER FREQUENCY (Hz)** | **250** | **354** | **500** | **707** | **1000** | **1410** | **2000** | **2830** | **4000** | **5660** |
|---|---|---|---|---|---|---|---|---|---|---|
| HP nominal cutoffs (Hz) | 250-354 | 354-500 | 500-707 | 707-1000 | 1000-1410 | 1410-2000 | 2000-2830 | 2830-4000 | 4000-5660 | 5660-8000 |
| **MEASURED CENTER FREQUENCY (Hz)** | **261** | **367** | **523** | **735** | **1043** | **1465** | **2091** | **2942** | **4172** | **5674** |
| Nominal Geometric Mean (Hz) | 297 | 421 | 595 | 841 | 1187 | 1679 | 2379 | 3365 | 4758 | 6729 |
| Lower bound @-20dB (Hz) | 221 | 311 | 439 | 620 | 885 | 1236 | 1770 | 2484 | 3504 | 4749 |
| Upper bound @-20dB (Hz) | 309 | 433 | 623 | 872 | 1228 | 1737 | 2471 | 3484 | 4968 | 6780 |
| Bandwidth @-20dB (Hz) | 88 | 122 | 184 | 253 | 343 | 501 | 701 | 1000 | 1464 | 2031 |
| Bandwidth (octave) | 0.49 | 0.48 | 0.51 | 0.49 | 0.47 | 0.49 | 0.48 | 0.49 | 0.50 | 0.51 |

AUTHORS' ORIGINAL MANUSCRIPT

### Supplemental Digital Content 2:

**SCHEMATIC EXAMPLE:**

**Calculation of $BW_{0.075}$ (bandwidths at 0.075 µV) from one subject's amplitude profile**

Subject #21's wave V amplitude profiles for 1-octave-wide derived responses to 2000-Hz NBchirp (red filled circles) and 2-1-2 (black open circles) stimuli.

Bandwidths of a subject's amplitude profiles determined for a wave V-V' amplitude of 0.075 µV on either side of the profile, shown schematically in this figure.

Blue dashed line = wave V amplitude of 0.075 µV

Thick red line: bandwidth for wave V to NBchirps

Thick black line: bandwidth for wave V to 2-1-2 stimuli

Calculated bandwidths* are 2606 Hz for NBchirps and 1789 Hz for 2-1-2 stimuli.

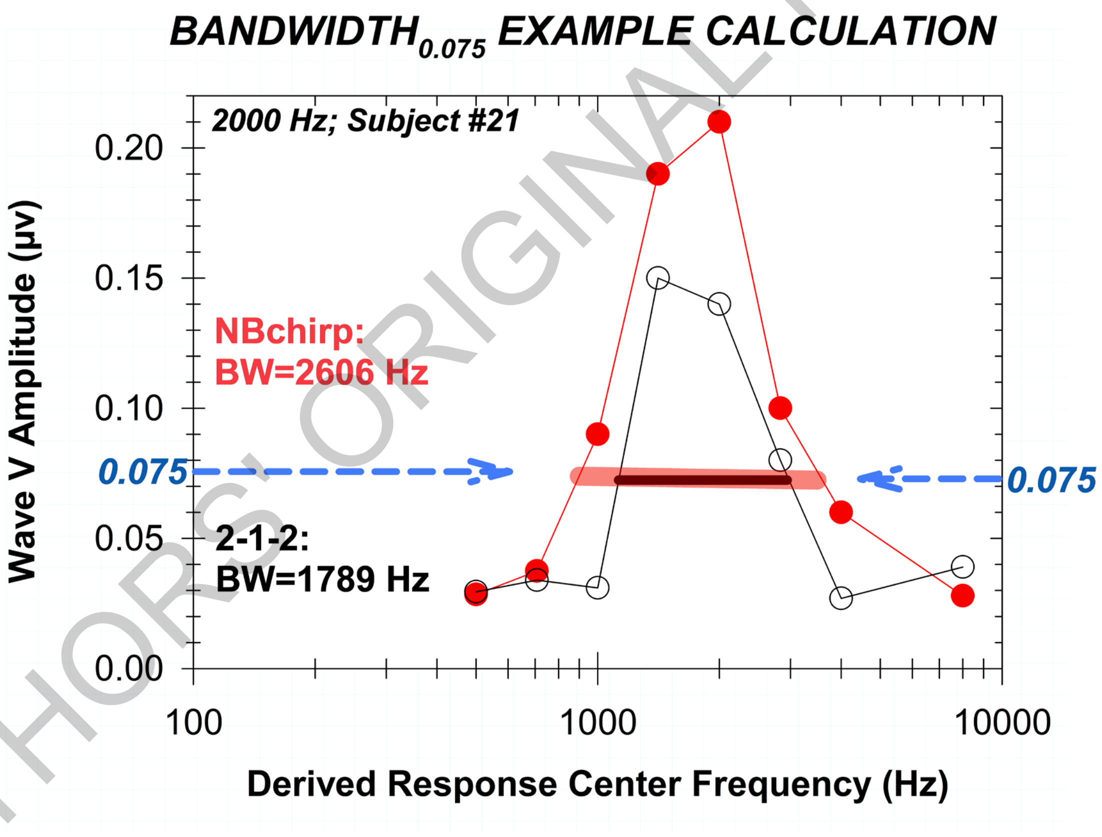


**Upper and lower frequencies were determined using linear regression between adjacent amplitudes.*

AUTHORS' ORIGINAL MANUSCRIPT

### SUPPLEMENTAL DIGITAL CONTENT 3

Individual (and grand average) waveforms for:

- 500 Hz: No-Mask; 1-octave DR500; ½-octave DR500
- 2000 Hz: No-Mask; 1-octave DR2000; ½-octave DR2000

AUTHORS' ORIGINAL MANUSCRIPT

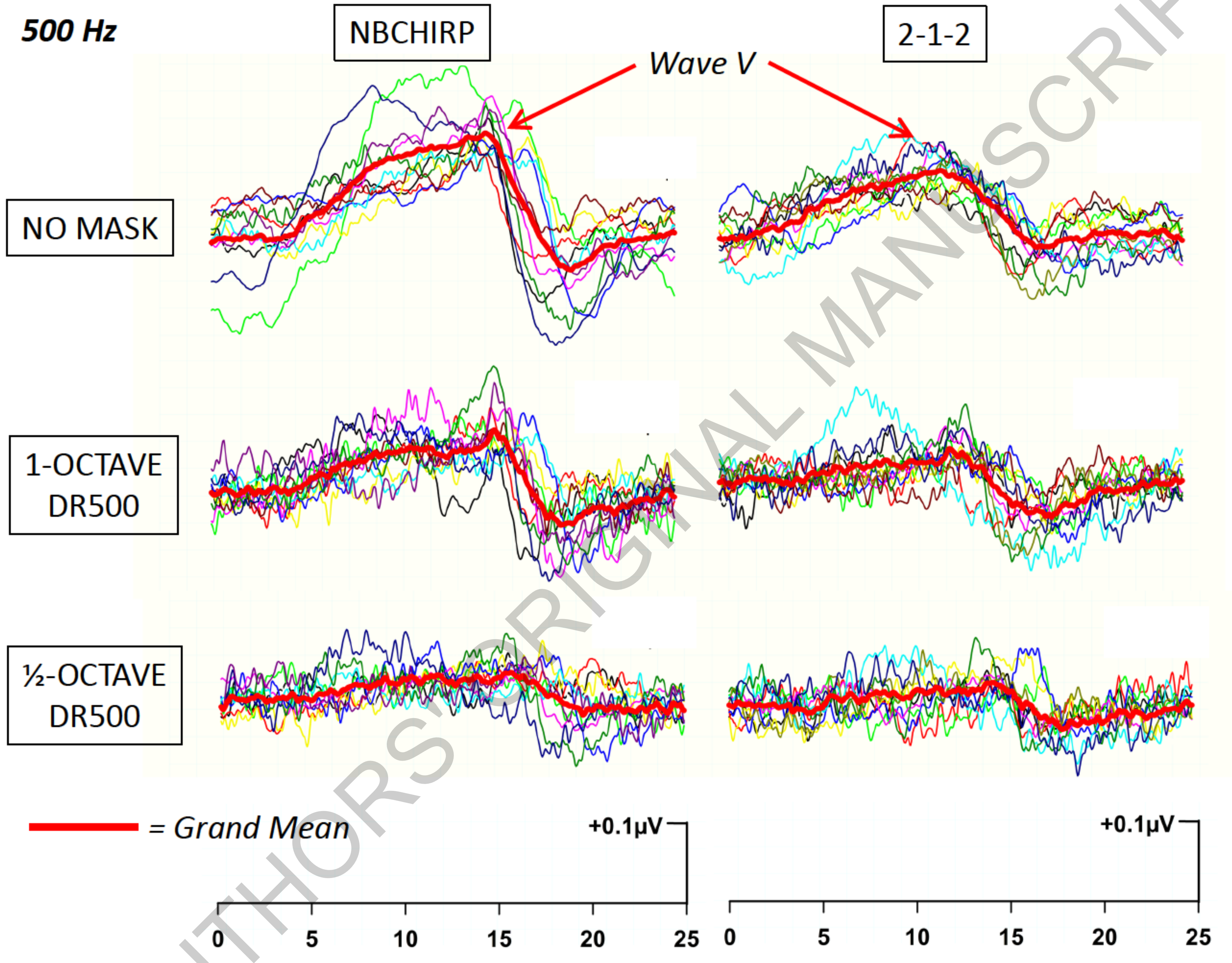

AUTHORS' ORIGINAL MANUSCRIPT

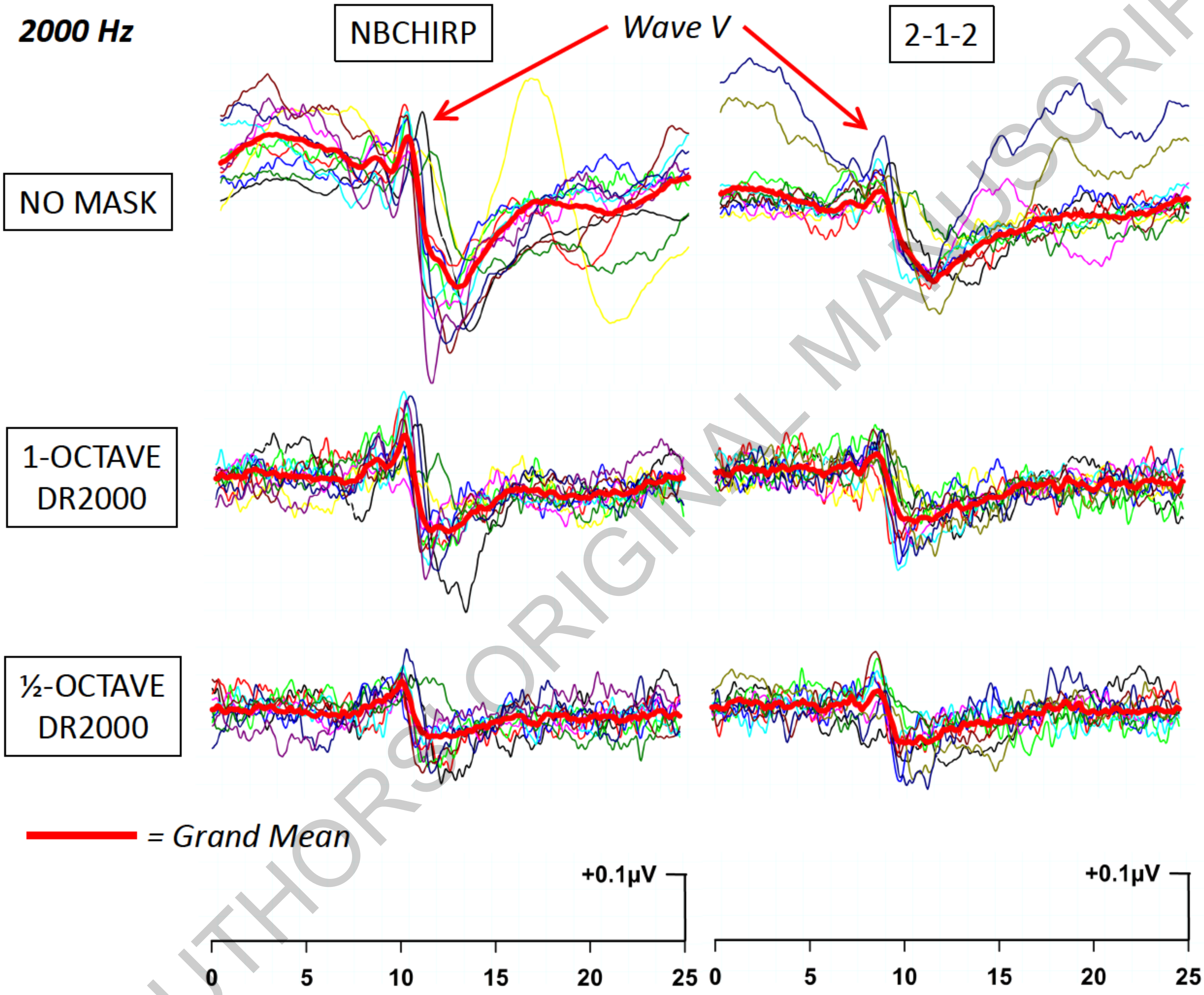

AUTHORS ORIGINAL MANUSCRIPT
